# Vaccine Effectiveness Against Long COVID in Children: A Report from the RECOVER EHR Cohort

**DOI:** 10.1101/2023.09.27.23296100

**Authors:** Hanieh Razzaghi, Christopher B. Forrest, Kathryn Hirabayashi, Qiong Wu, Andrea Allen, Suchitra Rao, Yong Chen, H. Timothy Bunnell, Elizabeth A. Chrischilles, Lindsay G. Cowell, Mollie R. Cummins, David A Hanauer, Miranda Higginbotham, Benjamin D. Horne, Carol R. Horowitz, Ravi Jhaveri, Susan Kim, Aaron Mishkin, Jennifer A Muszynski, Susanna Naggie, Nathan M. Pajor, Anuradha Paranjape, Hayden T Schwenk, Marion R. Sills, Yacob G. Tedla, David A. Williams, Charles Bailey, the RECOVER Consortium* A complete list of group members appears in the supplementary materials

**Affiliations:** Applied Clinical Research Center, Children’s Hospital of Philadelphia, Philadelphia, PA; Department of Pediatrics, Perelman School of Medicine, University of Pennsylvania, Philadelphia, PA; Department of Pediatrics, University of Colorado School of Medicine and Children’s Hospital Colorado, Aurora, CO; Department of Epidemiology, College of Public Health, University of Iowa, Iowa City, IA; Peter O’Donnell Jr. School of Public Health; Department of Immunology, School of Biomedical Sciences; UT Southwestern Medical Center; Dallas, TX; College of Nursing, University of Utah, Salt Lake City, UT; Department of Learning Health Sciences, University of Michigan, Ann Arbor, MI; Intermountain Heart Institute, Intermountain Health, Salt Lake City, UT; Division of Critical Care Medicine, Department of Pediatrics, Nationwide Children’s Hospital, Columbus, OH; Division of Infectious Diseases, Duke University School of Medicine; Duke Clinical Research Institute, Durham, NC; Department of Anesthesiology, University of Michigan, Ann Arbor, MI; Division of Infectious Diseases, Ann & Robert H. Lurie Children’s Hospital of Chicago, Chicago, IL; University of California, San Francisco, Division of Rheumatology, Benioff Children’s Hospital, San Francisco CA; Temple University Lewis Katz School of Medicine, Section of Infectious Diseases, Philadelphia, PA; Stanford School of Medicine, Division of Pediatric Infectious Diseases, Stanford, CA; Institute for Health Equity Research, Icahn School of Medicine at Mount Sinai, NYC, NY; Division of Epidemiology, Department of Medicine, Vanderbilt University Medical Center, Nashville, TN; Biomedical Research Informatics Center, Nemours Children’s Health, Wilmington, DE; OCHIN, Inc., Portland, OR; Division of Pulmonary Medicine, Cincinnati Children’s Hospital Medical Center and University of Cincinnati College of Medicine, Cincinnati, OH

**Author notes:** Address Correspondence to: Hanieh Razzaghi, Roberts Center for Pediatric Research, Room 11361, 2716 South Street, Philadelphia, PA 19146, [ ]. Disclosures: Dr. Mollie Cummins is employed by Doxy.me Inc., a commercial telemedicine platform provider. Dr. Benjamin Horne is a member of the advisory boards of Opsis Health and Lab Me Analytics, and a consultant to Pfizer (regarding clinical risk scores; funds paid to Intermountain). Dr. Susanna Naggie reports research grants from Gilead Sciences and AbbVie, scientific advisor/stock options from Vir Biotechnologies, consulting with no financial payment from Pardes Biosciences and Silverback Therapeutics, DSMB fees from Personal Health Insights, Inc, event adjudication committee fees from BMS/PRA outside the submitted work. Dr. Mishkin receives Grant support from Pfizer paid directly to Institution Advisory Board for Takeda. Dr. Jhaveri is a consultant for AstraZeneca, Seqirus, Dynavax, receives an editorial stipend from Elsevier and Pediatric Infectious Diseases Society. Dr. Rao reports prior grant support from GSK and Biofire and is a consultant for Sequiris. The remaining authors have no disclosures. Funding/Support: This research was funded by the National Institutes of Health (NIH) Agreement OT2HL161847-01 as part of the Researching COVID to Enhance Recovery (RECOVER) program of research. Disclaimer: The views and conclusions contained in this document are those of the authors and should not be interpreted as representing the official policies, either expressed or implied, of the NIH. Abbreviations: PCR – polymerase chain reaction; EHR – electronic health record; CI – 95% confidence intervals; ICD-10-CM – International Classification of Diseases, version 10, clinical modification; PMCA – Pediatric Medical Complexity Algorithm; US – United States of America; VE – vaccine effectiveness; VIS – vaccine information system.

## Abstract

**Objective:** Vaccination reduces the risk of acute COVID-19 in children, but it is less clear whether it protects against long COVID. We estimated vaccine effectiveness (VE) against long COVID in children aged 5-17 years.

**Methods:** This retrospective cohort study used data from 17 health systems in the RECOVER PCORnet electronic health record (EHR) Program for visits between vaccine availability, and October 29, 2022. Conditional logistic regression was used to estimate VE against long COVID with matching on age group (5-11, 12-17) and time period and adjustment for sex, ethnicity, health system, comorbidity burden, and pre-exposure health care utilization. We examined both probable (symptom-based) and diagnosed long COVID in the year following vaccination.

**Results:** The vaccination rate was 56% in the cohort of 1,037,936 children. The incidence of probable long COVID was 4.5% among patients with COVID-19, while diagnosed long COVID was 0.7%. Adjusted vaccine effectiveness within 12 months was 35.4% (95 CI 24.5 – 44.5) against probable long COVID and 41.7% (15.0 – 60.0) against diagnosed long COVID. VE was higher for adolescents 50.3% [36.3 – 61.0]) than children aged 5-11 (23.8% [4.9 – 39.0]). VE was higher at 6 months (61.4% [51.0 – 69.6]) but decreased to 10.6% (–26.8 – 37.0%) at 18-months.

**Discussion:** This large retrospective study shows a moderate protective effect of SARS-CoV-2 vaccination against long COVID. The effect is stronger in adolescents, who have higher risk of long COVID, and wanes over time. Understanding VE mechanism against long COVID requires more study, including EHR sources and prospective data.

**Article Summary:** Vaccination against COVID-19 has a protective effect against long COVID in children and adolescents. The effect wanes over time but remains significant at 12 months.

**What’s Known on This Subject:** Vaccines reduce the risk and severity of COVID-19 in children. There is evidence for reduced long COVID risk in adults who are vaccinated, but little information about similar effects for children and adolescents, who have distinct forms of long COVID.

**What This Study Adds:** Using electronic health records from US health systems, we examined large cohorts of vaccinated and unvaccinated patients <18 years old and show that vaccination against COVID-19 is associated with reduced risk of long COVID for at least 12 months.

**Contributors’ Statement:** Drs. Hanieh Razzaghi and Charles Bailey conceptualized and designed the study, supervised analyses, drafted the initial manuscript, and critically reviewed and revised the manuscript.

Drs. Christopher Forrest and Yong Chen designed the study and critically reviewed and revised the manuscript.

Ms. Kathryn Hirabayashi, Ms. Andrea Allen, and Dr. Qiong Wu conducted analyses, and critically reviewed and revised the manuscript.

Drs. Suchitra Rao, H Timothy Bunnell, Elizabeth A. Chrischilles, Lindsay G. Cowell, Mollie R. Cummins, David A. Hanauer, Benjamin D. Horne, Carol R. Horowitz, Ravi Jhaveri, Susan Kim, Aaron Mishkin, Jennifer A. Muszynski, Susanna Nagie, Nathan M. Pajor, Anuradha Paranjape, Hayden T. Schwenk, Marion R. Sills, Yacob G. Tedla, David A. Williams, and Ms. Miranda Higginbotham critically reviewed and revised the manuscript.

All authors approved the final manuscript as submitted and agree to be accountable for all aspects of the work.

**Authorship statement:** Authorship has been determined according to ICMJE recommendations.

## Introduction

SARS-CoV-2 has infected over 676 million people, and has been associated with 6.9 million deaths^1^. While severity has been lower in children than adults, statistics for the United States (US) indicate >2100 deaths in children aged <21 years with COVID-19^2^ and up to 2.1% of cases classified as severe disease^3^. Further, children may experience long-term health burdens after infection, collectively referred to as post-acute sequelae of SARS-CoV-2 (PASC) or “long COVID” ^4^ ^5,6^. These may represent effects of host response to the original infection in addition to or instead of direct pathogen effects^7–9^. Long COVID is heterogeneous and likely underdiagnosed, but symptoms such as brain fog, dyspnea, gastrointestinal dysfunction, generalized pain, and fatigue can cause significant burden for children, even after mild COVID-19^4,5,10,11^. Recent research has identified specific post-acute symptoms consistent with long COVID ^12^. Prevalence estimates vary widely^10,12–14^. Symptoms of long COVID have been reported less often and for shorter duration in children compared to adults ^11^, and higher rates have been reported for adolescents than younger children^13^. It is difficult to establish how much this results from differential reporting of symptoms at different ages, greater difficulty distinguishing long COVID from other childhood illnesses or effects of the pandemic (*e.g.* disruption of seasonal viral patterns, or of school progress)^13^. Despite the paucity of data about long COVID in children, developing mitigation strategies is an important concern not only for affected children but in societal discussions^15^ and for shaping public health policy^16–18^.

Vaccines provide the best opportunity for both reducing the risk of severe disease^19–24^ as well as short-term complications of acute infection. In adults, some reports^25–29^ identified a modest benefit associated with vaccination in preventing long COVID in adults, while others did not^30,31^. Little data are available on the effect of vaccination on long COVID in children; in one UK study including 6500 children with COVID-19, no association was seen^4^. However, to our knowledge, there are no studies assessing clinical data to address this question. Efforts to learn from clinical care at a large enough scale to identify underrepresented patterns provides an important complement to multicenter prospective cohorts. The NIH Researching COVID to Enhance Recovery (RECOVER) Initiative, which seeks to understand, treat, and prevent the post-acute sequelae of SARS-CoV-2 infection (PASC, or long COVID), is employing both strategies to better elucidate the condition, identify effective ways to treat patients, and support families impacted by long COVID. We report results from a large-scale, national collaboration of health systems from the PCORnet^®^ network contributing electronic health record (EHR) data as part of RECOVER’s real-world program^46^.

## Methods

### Data sources

We conducted a retrospective cohort study based on EHR data from 41 health systems for children with COVID-19, a diagnosis of acute respiratory illness, fever, dyspnea, or cough, or a SARS-CoV-2 vaccine. Data, extracted quarterly, were transformed to either the PCORnet (v6.0) or Observational Medical Outcomes Partnership (OMOP) (v5.3) data models, and forwarded to the RECOVER-PCORnet Coordinating Center. For this study, we used the s8 version of the data, collected in April 2023, which comprises 25,389,893 patients.

Institutional Review Board oversight was provided by the Biomedical Research Alliance of New York, protocol # 21-08-508-380.

### Study Sample

Patients were eligible if they had an in-person visit between vaccine eligibility and October 29, 2022, and were between 5 and <18 years at the date of vaccine eligibility. The date of SARS-CoV-2 vaccine eligibility was defined as January 01, 2021, for patients aged 12 – <18 years, and October 29, 2021, for patients aged 5 – <12 years, corresponding to availability of vaccines. A small number of patients were excluded for documented COVID-19 before 28 days after their first vaccine dose.

For vaccinated patients, cohort entry was the date of their first vaccine dose, while for unvaccinated patients, the date of an in-person visit after vaccine eligibility was selected randomly, resulting in matched distribution of entry dates between cohorts. In addition, we required 1 contact between 8 days and 2 years prior to cohort entry to assess baseline health characteristics. To determine long COVID outcomes, we also required at least 1 in-person visit between 28 and 179 days after cohort entry.

### Outcomes

The main outcomes were *diagnosed* or *probable* long COVID. A patient had *diagnosed* long COVID if they had two or more visits with diagnosis codes specific for long COVID; a single diagnosis code classified children as having *probable* long COVID. In addition, to account for incomplete availability or use of long-COVID-specific diagnosis codes, especially for patients with early signs of long COVID, we also classified a patient as having *probable* long COVID if they had COVID-19 (SARS-CoV-2 PCR or antigen positive or COVID-19-specific diagnosis codes) plus at least two long-COVID-compatible diagnoses 28-179 days after infection. These diagnoses were identified in prior work as post-acute associations with COVID-19^12,32^ (Supplemental Table S3).

Some results are reported for pre-omicron and omicron eras. This applies only to adolescents, as vaccines for younger children were not available substantially before the omicron era. To maintain consistency across analyses, November 29, 2021, the first day a younger child could have had a qualifying post-vaccine infection, was used as the dividing line between eras; at that point omicron was circulating in the US, and became the dominant strain a few weeks later. For each patient, the date of a SARS-CoV-2 infection following cohort entry, if one occurred, was used to place their results in either of the two eras; if the patient never developed COVID-19, the date of cohort entry was used.

### Exposure

The primary exposure was receipt of at least one dose of SARS-CoV-2 vaccine. Vaccine doses were ascertained using a vaccine code (CVX or RxNorm) designating an administered or patient-reported dose, a procedure code indicating SARS-CoV-2 vaccination, or a vaccine description containing the string “COVID” or “SARS”. Where available, data was retrieved from state/regional vaccine information systems via hospital operational procedures. Adequate vaccine data capture was defined as a site-reported vaccination rate at least 60% of the CDC-estimated rate^2^ for its coverage area, based on addresses of patients seen during the pandemic. Where dose counts were used, all recorded doses within 14 days of a base dose were considered a single dose.

### Demographic and clinical data

Age, sex, and ethnicity were taken from EHRs. Comorbidity data used the body system taxonomy of the Pediatric Medical Complexity Algorithm (PMCA)^14^. Diagnoses received up to 3 years before cohort entry were considered. To classify patients as complex-chronic, chronic, or no chronic condition, the “more conservative” version of the algorithm was applied.

### Main analysis

We matched vaccinated and unvaccinated patients on age group at cohort entry (5-11, 12-17) and time period (six-month cohort entry period). We limited long COVID observations to 12 months following cohort entry to account for waning of vaccine effectiveness (VE) over time.

### Sensitivity analyses

To evaluate potential impact of attrition due to matching, we performed a sensitivity analysis using inverse probability of treatment weighting that incorporated age group, time of cohort entry, sex, ethnicity, health system, number of baseline visits to the health system, PMCA-defined body system(s) affected by chronic illness, progressive disease, and malignancy. Further, we estimated VE after changing the window for outcomes to 6 months and 18 months post cohort entry and restricting the vaccinated cohort to those with at least 2 recorded doses.

### Cohort Comparison

For unadjusted analyses, follow-up time was computed from cohort entry until the end of observation. We calculated an incidence rate ratio (IRR) comparing vaccinated to unvaccinated groups and derived 95% CIs using the estimation of Wald^33^. Corresponding VE was computed as 100 × (1 IRR).

For adjusted analyses, we used conditional logistic regression to account for stratification. Covariates included sex, ethnicity, health system, baseline utilization (0, 1-5, 6-10, 11-24, 25-49, 50-99, or 100+ encounters in the 3 years prior to cohort entry), the number of PMCA body systems affected, and the presence of progressive or malignant condition. Since the prevalence of long COVID was low, IRR ≈ OR, so the corresponding VE was computed as 100 × (1-OR).

### Mediation Analyses

To understand the direct and indirect impact of vaccination on long COVID, we conducted causal mediation analyses, where the treatment was pre-infection vaccination, the mediator was SARS-CoV-2 infection, and the outcome was long COVID. We fit two logistic regression models, the first of treatment on the mediator, and the second of treatment and mediator on the outcome, adjusting for the same set of covariates. From these models, we estimated the effect and CIs of vaccination on long COVID independent of infection, and of vaccination on long COVID mediated by infection.

### Statistical Tools

Data analysis was performed using R^17^ version 4.0.2. Exact matching used the `matchIt` package ^34^ version 4.4.0, while IPTW were computed via `twang` ^35^ version 2.5. Conditional logistic regression used the `clogit()` algorithm implemented in the `survival` package ^36^ version 3.5. Significant effects were identified based on a threshold of *a*<0.05.

## Results

### Study Cohort

Immunization data from 17 health systems was sufficiently robust for inclusion. Cohort formation is described in Figure 1: the 5–11-year-old group comprised 480,498 children, while the 12-17 year-old group comprised 719,519 children/adolescents. Overall, 55% received at least one SARS-CoV-2 vaccine, and 84% of vaccinated children received two or more doses. Compared to unvaccinated children, vaccinated patients were older, had lower baseline healthcare utilization, and fewer chronic conditions. Girls and children who identified as Asian or Hispanic were more common in the vaccinated group, while those identifying as Black and White were less common (Table 1). Descriptive statistics for the matched cohorts are provided in Supplemental Table S1. Vaccinated patients had slightly fewer comorbidities and healthcare utilization than unvaccinated patients. The prevalence of probable long COVID was 0.3% in the cohort overall; for children with COVID-19 after cohort entry, prevalence of diagnosed long COVID was 0.8%, and adding probable cases raised the prevalence of long COVID to 4.5%.

**Figure 1:**
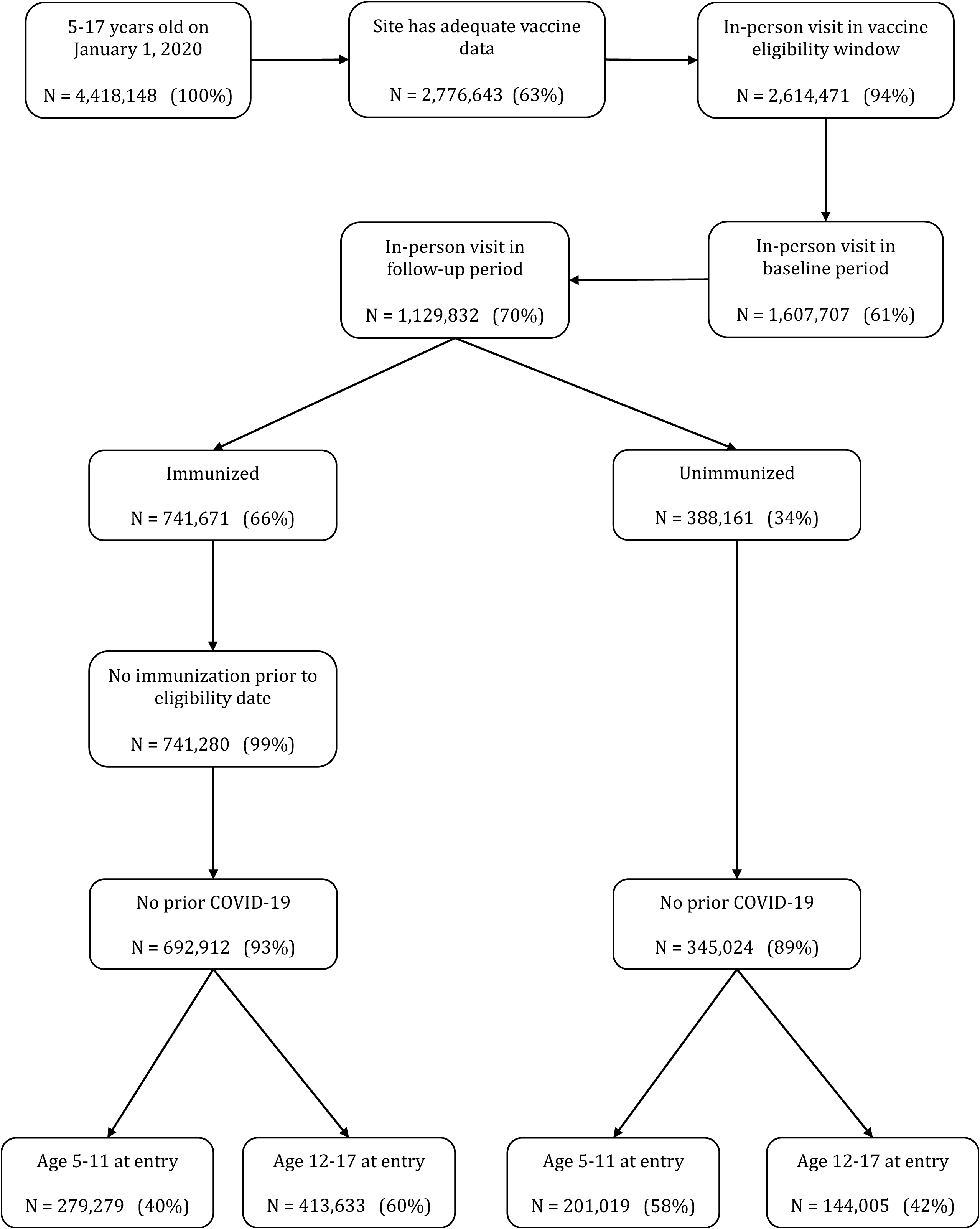
Study <u>Sample Selection</u>. Steps in the construction of the overall study sample are shown, including demographic and vaccine-related eligibility criteria. At each step, the number of children satisfying all criteria to that point is shown, as is the percentage of children in the previous step who were retained. ^a^ Vaccination rates ≥70% of CDC regional estimate

**Table 1.** Cohort Description for Full Cohort.

|  | **Vaccinated**<br>**(N = 692912)** | **Unvaccinated**<br>**(N = 345024)** | **Overall**<br>**(N = 1037936)** |
| --- | --- | --- | --- |
| **Race/Ethnicity** |  |  |  |
| Asian | 45267 (6.5%) | 8380 (2.4%) | 53647 (5.2%) |
| Black/AA (non-Hispanic) | 84642 (12.2%) | 63183 (18.3%) | 147825 (14.2%) |
| Hispanic | 168102 (24.3%) | 70976 (20.6%) | 239078 (23.0%) |
| Multiple/Other/<br>Missing/Unknown | 91494 (13.2%) | 36674 (10.6%) | 128168 (12.3%) |
| White (non-Hispanic) | 303407 (43.8%) | 165811 (48.1%) | 469218 (45.2%) |
| **Age (yr) at Entry<sup>a</sup>** |  |  |  |
| Mean (SD) | 12.4 (3.58) | 11.1 (3.66) | 11.9 (3.65) |
| Median [Min, Max] | 12.8 [5.02, 18.0] | 11.0 [5.01, 18.0] | 12.2 [5.01, 18.0] |
| **Age Group at Entry<sup>a</sup>** |  |  |  |
| 5-11 years | 279279 (40.3%) | 201019 (58.3%) | 480298 (46.3%) |
| 12-17 years | 413633 (59.7%) | 144005 (41.7%) | 557638 (53.7%) |
| **Sex** |  |  |  |
| Female/Missing | 353971 (51.1%) | 168787 (48.9%) | 522758 (50.4%) |
| Male | 338941 (48.9%) | 176237 (51.1%) | 515178 (49.6%) |
| **Time Period of Entry<sup>a</sup>** |  |  |  |
| Dec 2020 – May 2021 | 197799 (28.5%) | 38763 (11.2%) | 236562 (22.8%) |
| Jun – Nov 2021 | 319605 (46.1%) | 65391 (19.0%) | 384996 (37.1%) |
| Dec 2021 – May 2022 | 157720 (22.8%) | 135878 (39.4%) | 293598 (28.3%) |
| Jun – Nov 2022 | 17788 (2.6%) | 104992 (30.4%) | 122780 (11.8%) |
| **In-Person Visits in**<br>**Baseline Period** |  |  |  |
| 00 visits<sup>b</sup> | 44741 (6.5%) | 12236 (3.5%) | 56977 (5.5%) |
| 01 to 05 visits | 261683 (37.8%) | 122599 (35.5%) | 384282 (37.0%) |
| 06 to 10 visits | 168774 (24.4%) | 90751 (26.3%) | 259525 (25.0%) |
| 11 to 24 visits | 149470 (21.6%) | 83087 (24.1%) | 232557 (22.4%) |
| ≥25 visits | 68244 (9.9%) | 36351 (10.5%) | 104595 (10.1%) |
| **Chronic Disease Category<sup>c</sup>** |  |  |  |
| Non-complex Chronic | 63877 (9.2%) | 33448 (9.7%) | 97325 (9.4%) |
| Complex Chronic | 20983 (3.0%) | 12903 (3.7%) | 33886 (3.3%) |
| None | 608052 (87.8%) | 298673 (86.6%) | 906725 (87.4%) |
| **Progressive Condition<sup>c</sup>** |  |  |  |
| No | 654097 (94.4%) | 321720 (93.2%) | 975817 (94.0%) |
| Yes | 38815 (5.6%) | 23304 (6.8%) | 62119 (6.0%) |
| **Malignancy<sup>c</sup>** |  |  |  |
| No | 687759 (99.3%) | 341609 (99.0%) | 1029368 (99.2%) |
| Yes | 5153 (0.7%) | 3415 (1.0%) | 8568 (0.8%) |
| **Institution** |  |  |  |
| A | 23144 (3.3%) | 14769 (4.3%) | 37913 (3.7%) |
| B | 65697 (9.5%) | 29160 (8.5%) | 94857 (9.1%) |
| C | 142143 (20.5%) | 43523 (12.6%) | 185666 (17.9%) |
| D | 65036 (9.4%) | 46009 (13.3%) | 111045 (10.7%) |
| E | 15003 (2.2%) | 3495 (1.0%) | 18498 (1.8%) |
| F | 109402 (15.8%) | 55253 (16.0%) | 164655 (15.9%) |
| G | 17214 (2.5%) | 2057 (0.6%) | 19271 (1.9%) |
| H | 335 (0.0%) | 1696 (0.5%) | 2031 (0.2%) |
| I | 47910 (6.9%) | 33443 (9.7%) | 81353 (7.8%) |
| J | 26925 (3.9%) | 12387 (3.6%) | 39312 (3.8%) |
| K | 45811 (6.6%) | 17818 (5.2%) | 63629 (6.1%) |
| L | 39651 (5.7%) | 42678 (12.4%) | 82329 (7.9%) |
| M | 14732 (2.1%) | 11696 (3.4%) | 26428 (2.5%) |
| N | 2416 (0.3%) | 1371 (0.4%) | 3787 (0.4%) |
| O | 13352 (1.9%) | 6928 (2.0%) | 20280 (2.0%) |
| P | 23249 (3.4%) | 7076 (2.1%) | 30325 (2.9%) |
| Q | 14418 (2.1%) | 8021 (2.3%) | 22439 (2.2%) |
| R | 26474 (3.8%) | 7644 (2.2%) | 34118 (3.3%) |

<sup>a</sup> Vaccination date or visit date if unvaccinated
<sup>b</sup> Patients with baseline eligibility data based on administrative (*e.g.* telephone) visits
<sup>c</sup> Based on PMCA

### Effect of Vaccination on long COVID in Children and Adolescents

While the likelihood that a recorded vaccine is incorrectly attributed to a patient is very low, the risk that dose count is incomplete in reported data is well-described^37^, and discontinuation before a second dose is uncommon. It is therefore more accurate in this study design to include patients with evidence of vaccination, and, because any compensatory bias introduced thereby will tend to reduce estimates, more responsible to report results that do not overestimate vaccine effectiveness. We analyzed all children with evidence of vaccination prior to SARS-CoV-2 infection. Results were very similar across approaches: VE was 35% (95% CI 24– 45%) for the combined age groups (Table 2), with greater effect in adolescents than younger children. For children known to be completely vaccinated (*i.e.* 2+ doses) prior to SARS-CoV-2 infection, VE was 35% (95% CI 35– 53%) against probable long COVID within 12 months (Figure 2), with consistent findings within age strata, supporting the validity of our approach. For comparison, VE estimates against SARS-CoV-2 infection are shown in Supplemental Figure S1. We observed decreased likelihood of long COVID among boys and children with non-Hispanic Black or Asian ethnicity (Supplementary Table S2). The presence of chronic conditions had minimal effect, though likelihood of long COVID rose with increasing baseline utilization.

**Figure 2.**
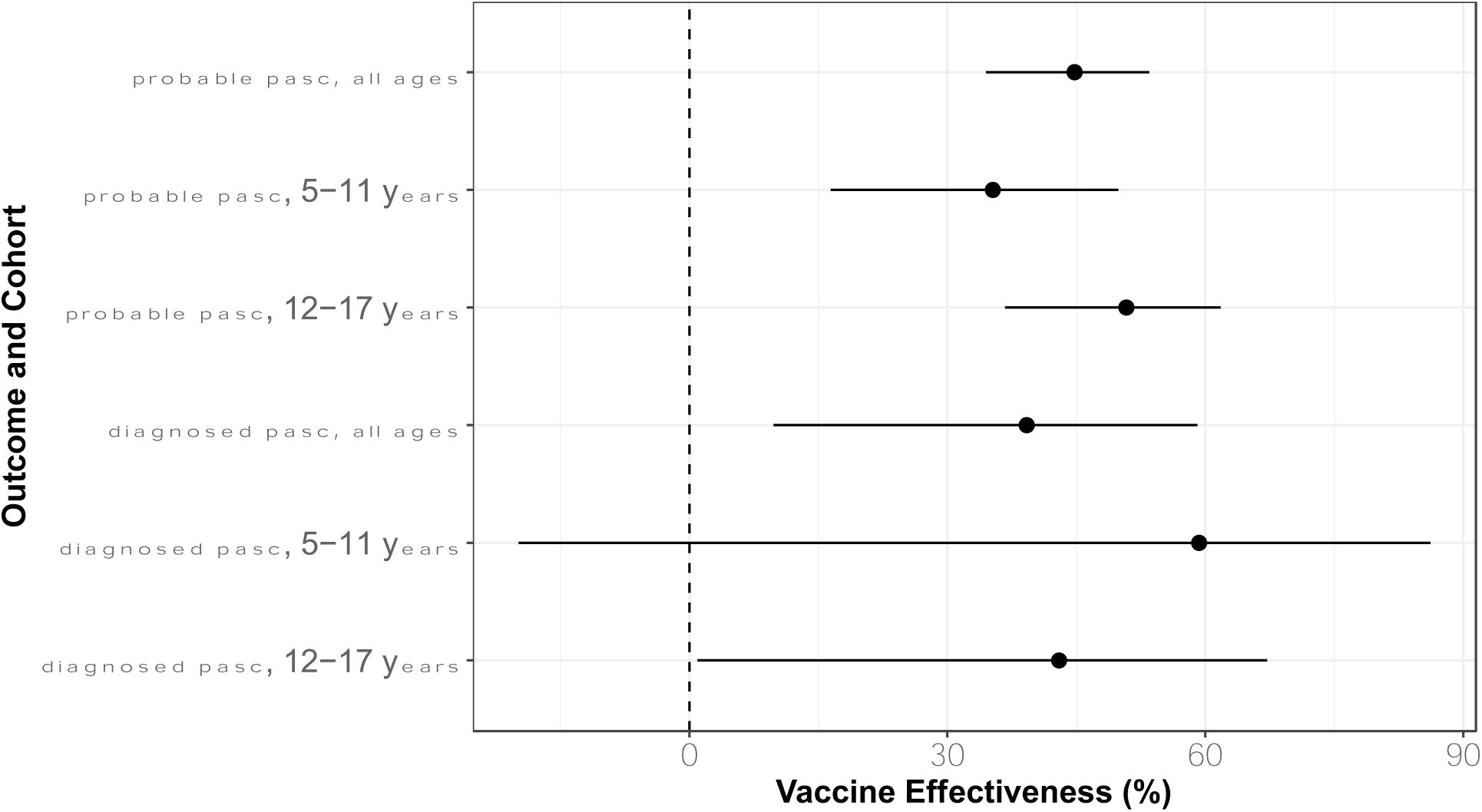
Vaccine effectiveness estimates for children receiving at least 2 doses of mRNA vaccine in their primary series. Analyses were otherwise structured identically to the main analysis.

**Table 2.** SARS-CoV-2 Vaccine Effectiveness Against Long COVID.

|  | N<sup>a</sup> | **Vaccine Effectiveness (%), 95% CI** |  |
| --- | --- | --- | --- |
|  |  | **Unadjusted** | **Adjusted<sup>b</sup>** |
| **Symptom-based or Diagnosed long COVID** |  |  |  |
| Child (5-11 y),<br>omicron | 753 | 19.7 (7.1 – 30.6) | 23.8 (4.9 – 39.0) |
| Adolescent (12-17 y),<br>all patients | 961 | 28.7 (18.8 – 37.4) | 50.3 (36.6 – 61.0) |
| Adolescent (12-17 y),<br>pre-omicron | 726 |  | 35.3 (4.8 – 56.0) |
| Adolescent (12-17y),<br>omicron |  |  | 58.7 (39.1 – 72.0) |
| All ages (5-17 y) | 1,669 | 25.3 (17.6 – 32.2) | 35.4 (24.5 – 44.5) |
| **Diagnosed long COVID only** |  |  |  |
| Child (5-11 y) | 82 | 16 (-32.7 – 46.9) | 48.2 (-.21 – 77.8) |
| Adolescent (12-17 y) | 194 | 41.7 (21.4 – 57.0) | 56.1 (25.8 – 74.0) |
| All ages (5-17 y) | 270 | 35.6 (17.4 – 50.0) | 41.7 (15.0 – 60.0) |

<sup>a</sup> Cases of long COVID. Note that values for 5-11 and 12-17 year age strata do not sum to the value for 5-17 as each stratum is drawn from sites with usable vaccine data for all ages in that stratum.
<sup>b</sup> Adjusted for sex, ethnicity, health system, baseline utilization in the 3 years prior to cohort entry, the number of body systems affected in the same interval, and the presence of a condition defined in the PMCA as either progressive or malignant

The protective effect of vaccination appeared to wane over time. We observed higher VE against long COVID within 6 months of vaccination than 12 months, while extending the observation period to 18 months revealed a further diminution (Figure 3). Protection was also seen in children who started vaccination after recuperating from COVID-19 (Supplemental Figure S2).

**Figure 3.**
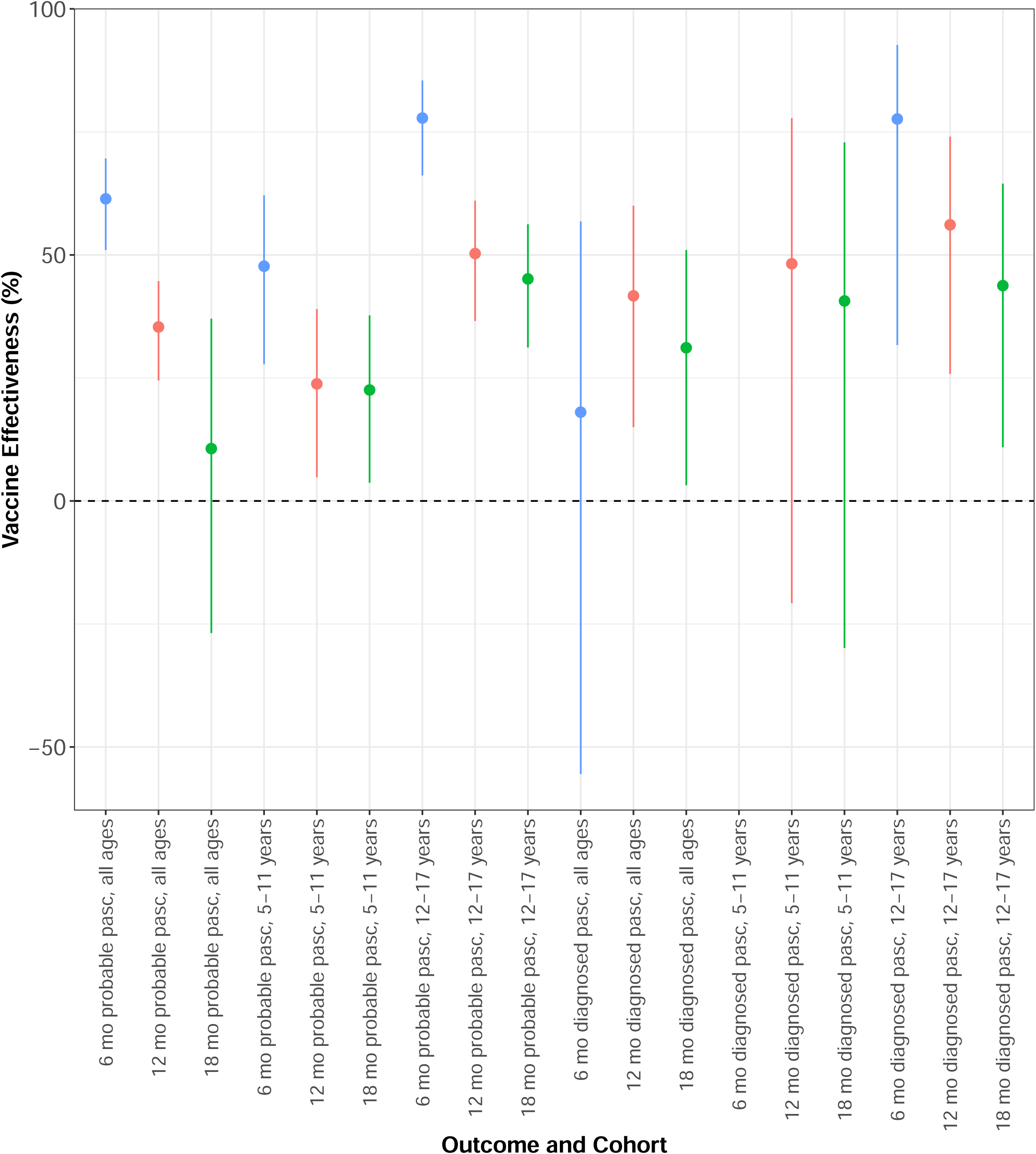
<u>Durability of vaccine effectiveness of SARS-CoV-2 vaccine against long COVID</u>. Results are shown for PASC occurring within 6, 12, or 18 months of vaccination.

One-to-one matching resulted in loss of 27% of unvaccinated and 64% of vaccinated patients due to the high overall vaccination rate. To assess the effect of this on our VE estimates, we used IPTW to adjust for multiple potential confounders without decreasing cohort size. Once again, a protective effect was seen, though with a smaller magnitude of 24% (95% CI 20 – 28%) for vaccination on long COVID up to a year later (Supplemental Figure S3).

To assess where in the path to long COVID the vaccine might have impact, we performed mediation analysis, estimating effect of vaccine on long COVID via change in COVID-19 risk (indirect effect) separately from other effects on long COVID (direct effect). For 5–11-year-olds, who became vaccine eligible just before the omicron variant became prevalent, the long COVID-associated OR for indirect effect on probable long COVID was 0.67 (95% CI 0.64 – 0.70), while for direct effect the OR was 1.34 (95% CI 1.13 – 1.59). For adolescents, results were similar: in the pre-omicron era the indirect effect OR was 0.46 (95% CI 0.45 – 0.48) and direct effect OR was 1.32 (95% CI 1.12 – 1.55); in the omicron era, the analogous ORs were 0.45 (95% CI 0.41 – 0.49) and 0.99 (95% CI 0.73 – 1.34). These results suggest that throughout the period of pediatric use, the effectiveness of vaccine against long COVID is closely tied to its effectiveness against the antecedent COVID-19 episode.

## Discussion

To our knowledge, this national-scale study, with a diverse cohort of over 1 million children, is the first to estimate the VE against long COVID in children. Our findings suggest that SARS-CoV-2 vaccine is associated with reduced risk of Long COVID in children and adolescents. The protective effect was greater among adolescents than younger children, reflecting activity against delta and omicron strains, while use for younger children is limited to the omicron era. Since diagnostic codes for long COVID (*e.g.* ICD-10-CM U09.9) are rare in children, we have examined both specific diagnosis as an outcome, and a broader definition based on the repeated presence from 1-6 months after infection of symptoms that occur more frequently following COVID-19 than in uninfected children^12,32^. The effect of vaccine is similar for both outcomes and is also seen in children vaccinated after an initial episode of COVID-19, suggesting that vaccination confers some benefit against future late effects. Mediation analysis suggests that the protective effect of vaccination may be primarily via reduction in the occurrence or severity of infection.

Our finding of a protective effect against long COVID in children are consistent with those observed in adults, while the VE estimates in our study are lower than studies assessing the short-term effect of vaccines in the one week to one month after vaccination ^29^. Other reasons are varied definitions of long COVID between these studies, and different methodological approaches. Our observed higher VE for adolescents is similar to studies examining acute infection ^38^. Further analyses are required to explore whether other factors contribute to this difference, such as long COVID symptoms being more frequently reported or documented in adolescents.

Waning effects of vaccine against long COVID are to be expected ^39^, similar to studies against acute infection ^40^ ^41^ ^42^. The reasons include reduced neutralization efficiency against newer strains and waning of pre-existing antibodies over time ^43^. It is also possible that long COVID starting several months after acute infection, or delayed diagnosis or recognition of long COVID, may contribute to this phenomenon. Further, waning effect may reflect the impact of successive infections rather than late-onset long COVID. This is difficult to assess analytically, given fewer COVID-19 episodes are being documented in the EHR during the omicron era. Evaluations of VE over time are needed to address whether these waning effects can be overcome through booster or annual vaccine doses.

VE studies using EHR data rely on accurate capture of clinical and vaccine data, with adjustment for important confounders to minimize potential bias ^44^ ^45^. Our focus on data quality, including comparison to the CDC’s county-level vaccination data, reduced our cohort size but yielded more reliable vaccination data. Our assessment of VE against acute SARS-CoV-2 was similar to VE estimates from other pediatric studies, further validating our findings and approach. Finally, we performed matching on age and time period to limit secular effects and conducted sensitivity analyses including inverse probability of treatment weighting, with similar findings.

Nonetheless, several challenges exist with the identification of long COVID in children using EHR data, which may confound assessment of our outcome. There has been low uptake of the long COVID (ICD10-CM U09.9) diagnosis code in pediatrics; it is more often seen in older children and associated with respiratory, cardiovascular, or fatigue subtypes of long COVID ^32^. For this reason, we supplemented our outcome definition with a symptom-based computable phenotype definition of long COVID we have developed, which includes features related to long COVID based on statistical association and chart review validation^12,32^. However, long COVID presentations are heterogeneous, and symptoms and conditions may overlap with other childhood illnesses, with the potential for misclassification of long COVID in our study. These limitations on classification accuracy are expected to affect both vaccinated and unvaccinated cohorts, similarly, so would not confound the comparison of the two.

There are other limitations that warrant discussion. First, our study relies on secondary use of EHR data, and is subject to bias due to differential access to and utilization of healthcare during the pandemic, as well as clinical practice variation across sites. We required both pre– and post-vaccine visits to establish a baseline of utilization, which may create bias toward sicker patients, but importantly we did not observe major imbalances in utilization rates between cohorts. Next, testing practices changed over time, with an increased reliance on home antigen testing in later phases of the pandemic, potentially leading to misclassification of symptom based long COVID. In addition, a large proportion of patients in our data with a long COVID diagnosis code do not have documentation in the EHR of the antecedent COVID-19 episode, which prevented use of severity as an adjustment to vaccine effect.

## Conclusion

Our results provide substantial evidence in a large and diverse cohort of children receiving health care for protective effect in children 5 years and older. This study adds to the growing body of knowledge about mitigating effect of vaccines on COVID-19, while demonstrating the need for further research utilizing a range of designs to examine the protective effect of these vaccines against subsequent strains and to help guide vaccine policy.

## Supporting information

RECOVER Consortium

## Data Availability

All data produced in the present study are available upon reasonable request to the authors

https://redcap.chop.edu/surveys/?s=88WJ8WAFPJ

## Acknowledgments

We are grateful to the health systems, clinicians, and patients at member sites without whose trust research of this type would not be possible. We are also grateful to clinical data teams at each RECOVER site involved in data extraction and integration, as well as colleagues at the RECOVER Coordinating Center, particularly Susan Hague, for maintaining the RECOVER data infrastructure. Additionally, we would like to thank the National Community Engagement Group (NCEG), all patient, caregiver and community Representatives, and all the participants enrolled in the RECOVER Initiative

