## Supplement for "Vaccine Effectiveness Against Long COVID in Children: A Report from the RECOVER EHR Cohort"

**Supplemental Table S1**. Cohort Description for Matched Cohort

|  | Vaccinated  (N = 692912) | Unvaccinated (N = 345024) | Overall (N = 1037936) |
| --- | --- | --- | --- |
| Race/Ethnicity |  |  |  |
| Asian | 15174 (6.3%) | 4845 (2.0%) | 20019 (4.1%) |
| Black/AA (non-Hispanic) | 45087 (18.7%) | 46096 (19.1%) | 91183 (18.9%) |
| Hispanic | 62390 (25.9%) | 45939 (19.0%) | 108329 (22.5%) |
| Multiple/Other/  Missing/Unknown | 28095 (11.6%) | 24012 (10.0%) | 52107 (10.8%) |
| White (non-Hispanic) | 90473 (37.5%) | 120327 (49.9%) | 210800 (43.7%) |
| Age (yr) at Entry^a^ |  |  |  |
| Mean (SD) | 11.4 (3.52) | 11.3 (3.75) | 11.4 (3.64) |
| Median [Min, Max] | 11.5 [5.02, 18.0] | 11.5 [5.01, 18.0] | 11.5 [5.01, 18.0] |
| Age Group at Entry^a^ |  |  |  |
| 5-11 years | 129961 (53.9%) | 129961 (53.9%) | 259922 (53.9%) |
| 12-17 years | 111258 (46.1%) | 111258 (46.1%) | 222516 (46.1%) |
| Sex |  |  |  |
| Female/Missing | 119957 (49.7%) | 118595 (49.2%) | 238552 (49.4%) |
| Male | 121262 (50.3%) | 122624 (50.8%) | 243886 (50.6%) |
| Time Period of Entry^a^ |  |  |  |
| Dec 2020 – May 2021 | 33310 (13.8%) | 33310 (13.8%) | 66620 (13.8%) |
| Jun – Nov 2021 | 62076 (25.7%) | 62076 (25.7%) | 124152 (25.7%) |
| Dec 2021 – May 2022 | 128681 (53.3%) | 128681 (53.3%) | 257362 (53.3%) |
| Jun – Nov 2022 | 17152 (7.1%) | 17152 (7.1%) | 34304 (7.1%) |
| In-Person Visits in Baseline Period |  |  |  |
| 00 visits^b^ | 9279 (3.8%) | 7479 (3.1%) | 16758 (3.5%) |
| 01 to 05 visits | 84954 (35.2%) | 79177 (32.8%) | 164131 (34.0%) |
| 06 to 10 visits | 63793 (26.4%) | 64501 (26.7%) | 128294 (26.6%) |
| 11 to 24 visits | 58504 (24.3%) | 62194 (25.8%) | 120698 (25.0%) |
| ≥25 visits | 24689 (10.2%) | 27868 (11.6%) | 52557 (10.9%) |
| Chronic Disease Category^c^ |  |  |  |
| Chronic, non-complex | 25524 (10.6%) | 24634 (10.2%) | 50158 (10.4%) |
| Complex Chronic | 8715 (3.6%) | 9479 (3.9%) | 18194 (3.8%) |
| None | 206980 (85.8%) | 207106 (85.9%) | 414086 (85.8%) |
| Progressive Condition^c^ |  |  |  |
| No | 226813 (94.0%) | 224343 (93.0%) | 451156 (93.5%) |
| Yes | 14406 (6.0%) | 16876 (7.0%) | 31282 (6.5%) |
| Malignancy^c^ |  |  |  |
| No | 239238 (99.2%) | 238714 (99.0%) | 477952 (99.1%) |
| Yes | 1981 (0.8%) | 2505 (1.0%) | 4486 (0.9%) |
| Institution |  |  |  |
| A | 4820 (2.0%) | 9988 (4.1%) | 14808 (3.1%) |
| B | 3225 (1.3%) | 20096 (8.3%) | 23321 (4.8%) |
| C | 43017 (17.8%) | 27536 (11.4%) | 70553 (14.6%) |
| D | 23634 (9.8%) | 32856 (13.6%) | 56490 (11.7%) |
| E | 9452 (3.9%) | 30732 (12.7%) | 40184 (8.3%) |
| F | 92048 (38.2%) | 50430 (20.9%) | 142478 (29.5%) |
| G | 3289 (1.4%) | 1474 (0.6%) | 4763 (1.0%) |
| H | 92048 (38.2%) | 50430 (20.9%) | 142478 (29.5%) |
| I | 13045 (5.4%) | 23479 (9.7%) | 36524 (7.6%) |
| J | 3382 (1.4%) | 5173 (2.1%) | 8555 (1.8%) |
| K | 19781 (8.2%) | 12611 (5.2%) | 32392 (6.7%) |
| L | 9452 (3.9%) | 30732 (12.7%) | 40184 (8.3%) |
| M | 19781 (8.2%) | 12611 (5.2%) | 32392 (6.7%) |
| N | 2878 (1.2%) | 2391 (1.0%) | 5269 (1.1%) |
| O | 522 (0.2%) | 4920 (2.0%) | 5442 (1.1%) |
| P | 18127 (7.5%) | 6345 (2.6%) | 24472 (5.1%) |
| Q | 2191 (0.9%) | 5682 (2.4%) | 7873 (1.6%) |
| R | 1808 (0.7%) | 7506 (3.1%) | 9314 (1.9%) |

^a^ Vaccination date or visit date if unvaccinated

^b^ Patients with baseline eligibility data based on administrative (*e.g.* telephone) visits

^c^ Based on PMCA

**Supplemental Figure S1**. Vaccine effective estimates against SARS-CoV-2 infection in the 12 months following vaccination. Infection was defined as a positive viral test (NAA or antigen) or appearance of a specific diagnosis for COVID-19. To allow for more direct comparison with long COVID analyses, the dividing line of November 30, 2021 between the pre-omicron and omicron eras was used in this analysis as well.

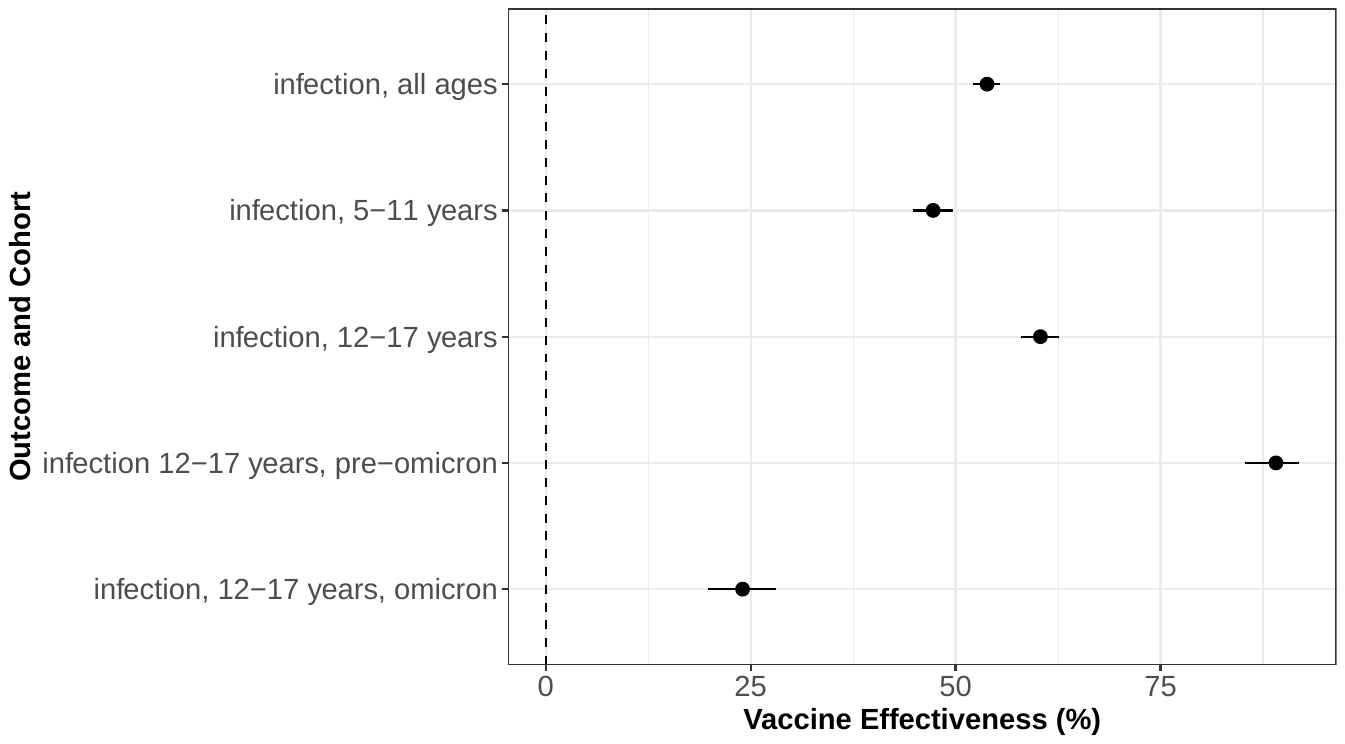

**Supplemental Table S2**. Covariate effect estimates in multivariable regression for the main analysis.

| **Variable** | **Odds Ratio** | **95% CI** | **P value** |
| --- | --- | --- | --- |
| **Immunization Status** | | | |
| Immunized prior to infection | 0.65 | 0.55 - 0.76 | 0 |
| Not immunized prior to infection | *Referent* | | |
| **Site** | | | |
| Site 1 | 1.25 | 0.82 - 1.91 | 0.3 |
| Site 2 | 1.36 | 0.99 - 1.87 | 0.06 |
| Site 3 | 1.49 | 0.8 - 2.76 | 0.21 |
| Site 4 | 0.09 | 0.04 - 0.22 | 0 |
| Site 5 | 2.7 | 1.1 - 6.62 | 0.03 |
| Site 6 | 1.69 | 0.95 - 3.02 | 0.08 |
| Site 7 | 0.73 | 0.33 - 1.63 | 0.44 |
| Site 8 | 0.54 | 0.12 - 2.49 | 0.43 |
| Site 9 | 1.05 | 0.65 - 1.67 | 0.85 |
| Site 10 | 0.93 | 0.63 - 1.36 | 0.69 |
| Site 11 | 1.47 | 1 - 2.16 | 0.05 |
| Site 12 | 1.3 | 0.67 - 2.49 | 0.44 |
| Site 13 | 0.88 | 0.28 - 2.78 | 0.83 |
| Site 14 | 2.78 | 1.5 - 5.17 | 0 |
| Site 15 | *Referent* | | |
| **Race/Ethnicity** | | | |
| Asian/Pacific Islander | 0.47 | 0.27 - 0.81 | 0.01 |
| Non-Hispanic Black or African American | 0.67 | 0.53 - 0.84 | 0 |
| Hispanic | 0.93 | 0.74 - 1.16 | 0.5 |
| Other or Unknown | 1.07 | 0.79 - 1.45 | 0.65 |
| Non-Hispanic White | *Referent* | | |
| **Sex** | | | |
| Female | 1.44 | 1.22 - 1.71 | 0 |
| Male | *Referent* | | |
| **Visit Utilization** | | | |
| 1-5 visits | 1.43 | 0.84 - 2.42 | 0.18 |
| 6-10 visits | 1.77 | 1.03 - 3.05 | 0.04 |
| 11-24 visits | 2.8 | 1.63 - 4.78 | 0 |
| 25-99 visits | 5.82 | 3.27 - 10.36 | 0 |
| 50-99 visits | 12.28 | 6.27 - 24.07 | 0 |
| 100+ visits | 15.62 | 6.69 - 36.47 | 0 |
| 0 visits | *Referent* | | |
| **Medical Complexity** | | | |
| PMCA number of body systems | 1.03 | 0.96 - 1.11 | 0.43 |
| PMCA progressive conditions | 1.49 | 1.05 - 2.1 | 0.02 |
| PMCA malignancy code | 1.79 | 0.93 - 3.47 | 0.08 |

**Supplemental Figure S2**. Vaccine effectiveness against long COVID after documented COVID-19. Documentation of COVID-19 by viral testing of specific diagnosis prior to cohort entry was required; imputed dates of infection were not considered in this step. Results for diagnosed PASC in the 5-11 year old group are not shown due to the small number of cases.

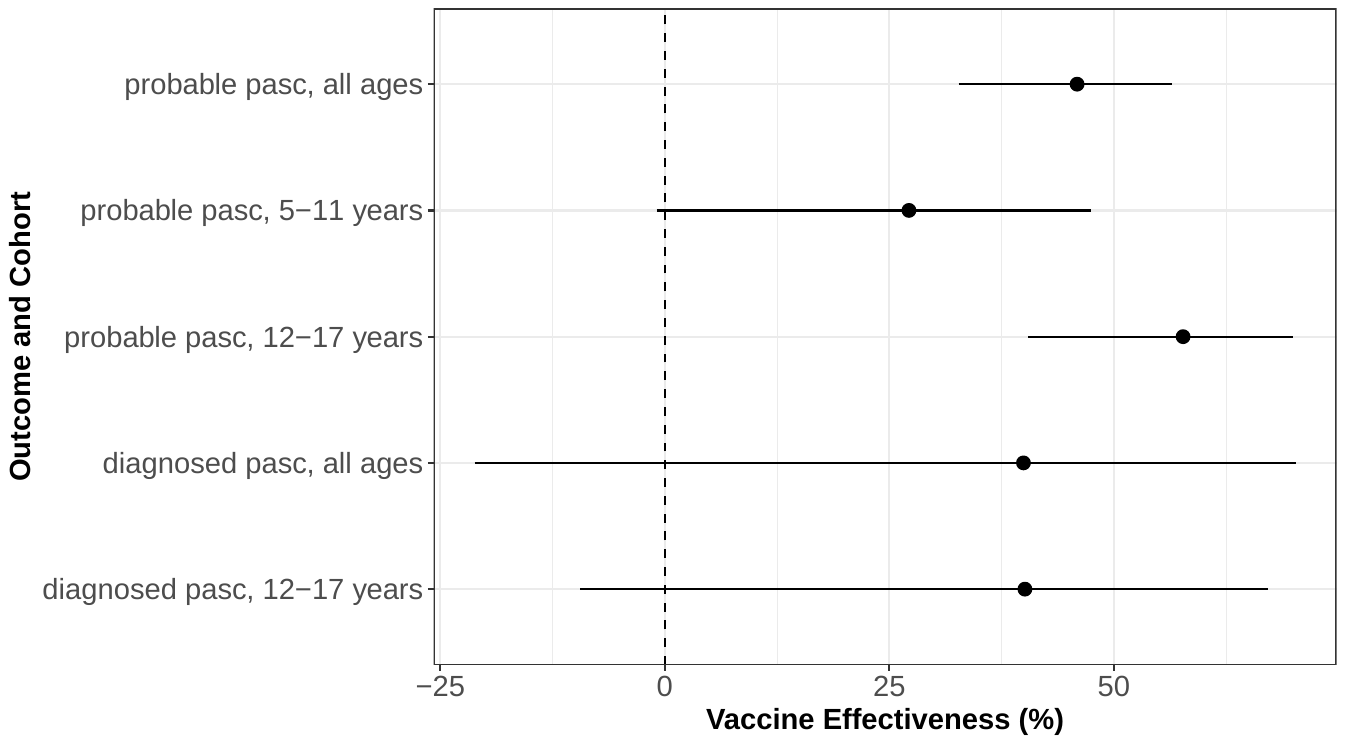

**Supplemental Figure S3**. Vaccine effectiveness against long COVID using inverse probability of treatment weighting (IPTW). Weights were computed using age stratum (5-11, 12-17), six month period of cohort entry, sex, ethnicity, health system, presence of a chronic condition at baseline in each body system defined in the PMCA taxonomy, and number of visits in the baseline period.

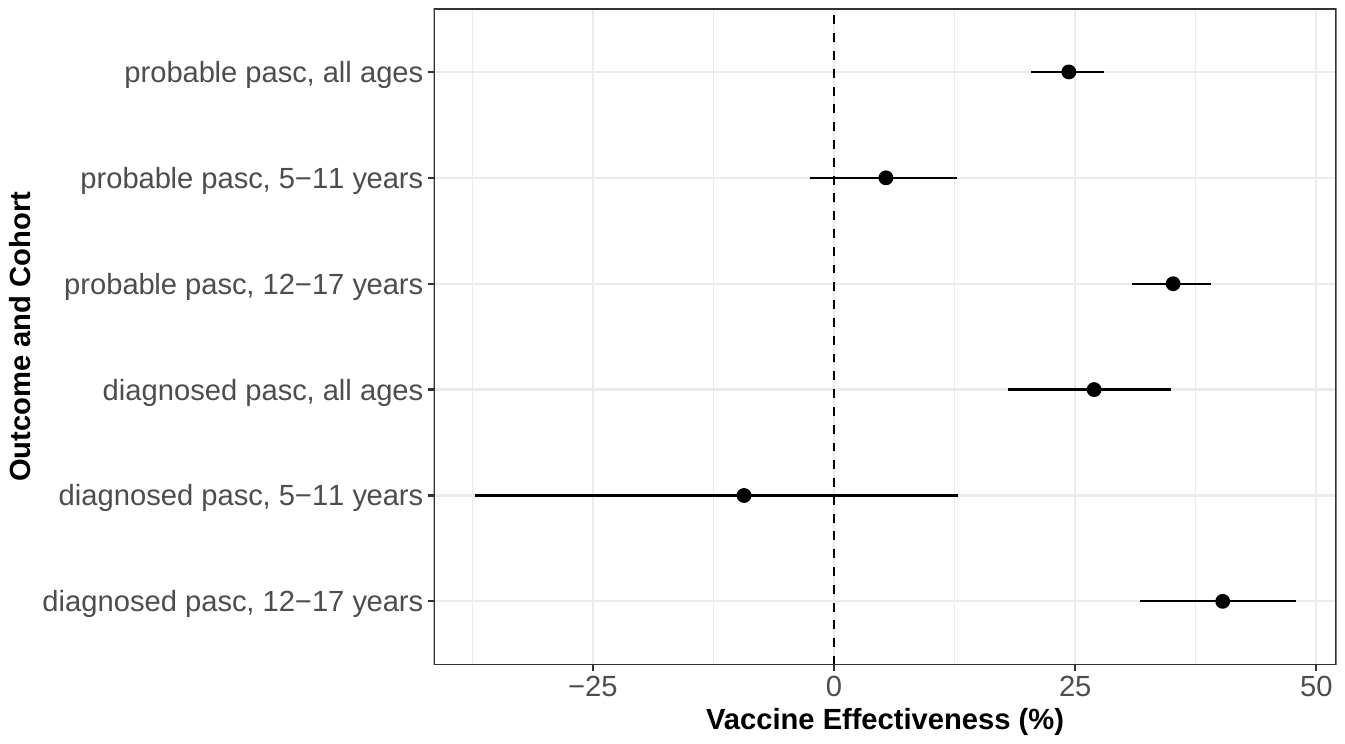

### Evaluating breakthrough infection

Understanding whether the development of COVID-19 after vaccination (“breakthrough infection”) provides information about risk for long COVID is important for several reasons. First, it may help to identify a group of patients who can benefit from increased screening, or early access to treatments designed to prevent long COVID or subsequent episodes of COVID-19, as these become available. Second, this pattern may be informative about immunologic status in ways that help us to understand potential mechanisms for long COVID. Third, it may help inform consideration of whether these patients should be considered a priority group when new vaccines, more effective against more recent strains of SARS-CoV-2, become available.

However, studying this question is complicated for many reasons. Long COVID is necessarily dependent on having had COVID-19, but the length of time after infection that a person is at risk to develop long COVID is unknown. In some cases the episode of COVID-19 may not be recognized or documented until long COVID occurs. This is particularly true in the time since omicron and subsequent strains of SARS-CoV-2 became prevalent, both because acute illness is milder than earlier in the pandemic, and because the wide use of over-the-counter testing at home or elsewhere means that many times COVID-19 is not documented in the medical record. In addition, it has become more likely over time that an individual has had more than one episode of COVID-19; if they develop long COVID it may be unclear to which episode(s) of COVID-19 it is most related.

Conversely, it is very difficult, for some of the same reasons, to undertake an experimental study of this problem. Documentation of prior COVID-19 episodes is challenging for any study, particularly with higher seroprevalence in the population making it difficult to identify individuals with no evidence of SARS-CoV-2 exposure. Randomized clinical trials addressing this question will be difficult because of the changing frequency and manifestations of long COVID, though prospective cohort studies will provide valuable information. Randomization to non-vaccination also presents ethical dilemmas given the vaccine’s known effectiveness in preventing severe acute illness.

While aware of these complexities, we have undertaken an exploratory analysis of breakthrough infection in this large, longitudinal cohort of children and adolescents. We utilized the same cohort definitions as the main analysis, but in both arms of the comparison required evidence of SARS-CoV-2 infection, as described in Methods, at least 28 days following cohort entry, and long COVID outcome was assessed starting 28 days after the date of breakthrough infection, but occurring no more than 12 months from cohort entry.

In this analysis, we observed a small positive association between vaccination and long COVID, with an OR of 1.32 (95% CI 1.18 – 1.49) for diagnosed or probable long COVID in the combined age group (Supplemental Figure S4). For diagnosed long COVID only, the odds ratio was 1.78 (95% CI 1.35 – 2.34). Estimates did not differ strongly across age groups, though the smaller number of cases in younger children results in wider confidence intervals.

**Supplemental Figure S4**. Occurrence of long COVID following breakthrough SARS-CoV-2 infection.

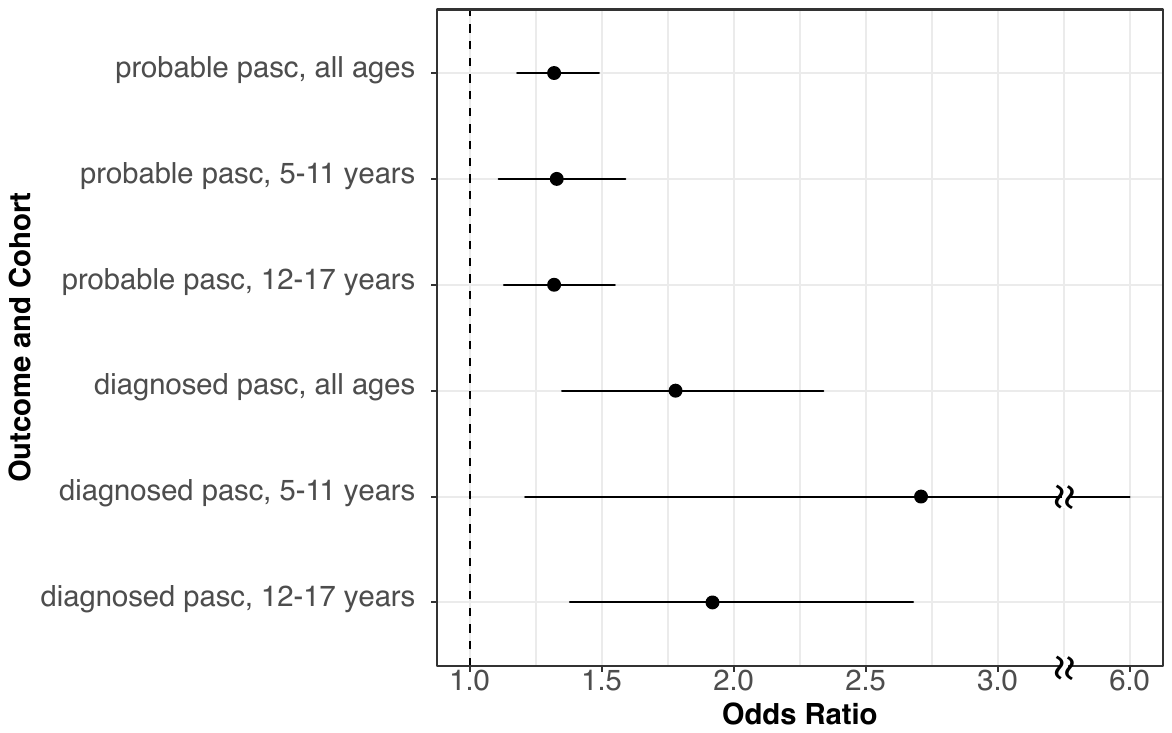

The observed association could result from any of several mechanisms; it is highly unlikely that vaccination is causally related to long COVID when the opposite is observed in the larger analyses. The occurrence of clinically evident COVID-19 despite vaccination may be a marker of some intrinsic aspect of host-pathogen interaction that bears on long COVID risk as well as infection itself. Alternatively, characteristics of host immune response may be modified in vaccinated patients in ways that affect autoimmune manifestations of long COVID. It is of course also possible that the association results from unmeasured confounding. As the main analysis showed, neither vaccination nor the utilization requirements that were necessary to assess other study variables were themselves causes of increased long COVID in vaccinated patients; confounding here would need to depend on the breakthrough infection event. Because we required the same level of recording for COVID-19 in both arms of the analysis, such confounding would need to differentially affect the likelihood that long COVID symptoms would be recorded for a vaccinated patient, separately from identifying breakthrough COVID-19. This is difficult to attribute to any of the health care or EHR processes used here.. We considered carefully whether the additional constraint of breakthrough infection was itself a collider. In order to satisfy that definition it must be independently affected by the exposure (vaccination) and outcome (long COVID). While the former is the case, the latter cannot be: long COVID is a consequence of COVID-19, and cannot affect the antecedent infection. Therefore breakthrough infection does not per se create collider bias. Nonetheless, this does not eliminate other potential biases, including ascertainment bias due to differences in patient or clinician behavior across cohorts. For example, it is possible that in unvaccinated patients the threshold to report COVID-19-like illness is lower than in vaccinated patients. This might result in identifying more mild breakthrough COVID-19 episode in unvaccinated patients, and if acute severity is associated with later development of long COVID, an artifactual association with vaccination may be seen.

Some risk of unmeasured confounding or collision is inherent in observational studies, or indeed in any study design. Therefore findings such as this limited association must be evaluated carefully, particularly when plausible underlying mechanisms are not well identified. Further study will be required to support or refute this result, both through similar studies in differing populations and through clinical and laboratory experimental studies.

**Supplemental Table S3**. Diagnosis codes used to assess for symptom-based (probable) long COVID. Diagnoses are shown as ICD-10-CM codes as well as concept IDs in the Observational Health Data Sciences and Informatics (OHDSI) collaborative vocabulary. Codes are grouped into subphenotypes grouping related pathophysiology.

| **Concept ID** | **Name** | **ICD-10-CM Code** | **Subphenotype** |
| --- | --- | --- | --- |
| 45602005 | Left lower quadrant rebound abdominal tenderness | R10.824 | abdominal_pain |
| 45553715 | Left lower quadrant pain | R10.32 | abdominal_pain |
| 45597170 | Right lower quadrant rebound abdominal tenderness | R10.823 | abdominal_pain |
| 45582694 | Right lower quadrant pain | R10.31 | abdominal_pain |
| 45568114 | Unspecified abdominal pain | R10.9 | abdominal_pain |
| 45577782 | Generalized abdominal tenderness | R10.817 | abdominal_pain |
| 45597169 | Left upper quadrant rebound abdominal tenderness | R10.822 | abdominal_pain |
| 1572201 | Other abdominal pain | R10.8 | abdominal_pain |
| 45573010 | Abdominal tenderness, unspecified site | R10.819 | abdominal_pain |
| 45544130 | Right lower quadrant abdominal tenderness | R10.813 | abdominal_pain |
| 1572198 | Abdominal and pelvic pain | R10 | abdominal_pain |
| 45582695 | Right upper quadrant rebound abdominal tenderness | R10.821 | abdominal_pain |
| 45602006 | Epigastric rebound abdominal tenderness | R10.826 | abdominal_pain |
| 45592408 | Abdominal tenderness | R10.81 | abdominal_pain |
| 45577783 | Periumbilic rebound abdominal tenderness | R10.825 | abdominal_pain |
| 45606795 | Left lower quadrant abdominal tenderness | R10.814 | abdominal_pain |
| 45548948 | Generalized rebound abdominal tenderness | R10.827 | abdominal_pain |
| 45602004 | Periumbilic abdominal tenderness | R10.815 | abdominal_pain |
| 45592407 | Periumbilical pain | R10.33 | abdominal_pain |
| 1572202 | Rebound abdominal tenderness | R10.82 | abdominal_pain |
| 45553716 | Rebound abdominal tenderness, unspecified site | R10.829 | abdominal_pain |
| 35211289 | Acute abdomen | R10.0 | abdominal_pain |
| 45573009 | Right upper quadrant abdominal tenderness | R10.811 | abdominal_pain |
| 45563290 | Lower abdominal pain, unspecified | R10.30 | abdominal_pain |
| 45558455 | Generalized abdominal pain | R10.84 | abdominal_pain |
| 45602007 | Colic | R10.83 | abdominal_pain |
| 45577781 | Upper abdominal pain, unspecified | R10.10 | abdominal_pain |
| 45558454 | Epigastric pain | R10.13 | abdominal_pain |
| 45534427 | Left upper quadrant abdominal tenderness | R10.812 | abdominal_pain |
| 45597168 | Left upper quadrant pain | R10.12 | abdominal_pain |
| 45534428 | Epigastric abdominal tenderness | R10.816 | abdominal_pain |
| 45568112 | Right upper quadrant pain | R10.11 | abdominal_pain |
| 45552530 | Abdominal migraine, not intractable | G43.D0 | abdominal_pain |
| 45533140 | Abdominal migraine, intractable | G43.D1 | abdominal_pain |
| 35211421 | Nonspecific elevation of levels of transaminase and lactic acid dehydrogenase [LDH] | R74.0 | abnormal_liver_enzymes |
| 725460 | Elevation of levels of liver transaminase levels | R74.01 | abnormal_liver_enzymes |
| 725461 | Elevation of levels of lactic acid dehydrogenase [LDH] | R74.02 | abnormal_liver_enzymes |
| 35211524 | Abnormal results of liver function studies | R94.5 | abnormal_liver_enzymes |
| 1571485 | Acute kidney failure | N17 | acute_kidney_injury |
| 35209269 | Acute kidney failure with tubular necrosis | N17.0 | acute_kidney_injury |
| 35209272 | Other acute kidney failure | N17.8 | acute_kidney_injury |
| 35209271 | Acute kidney failure with medullary necrosis | N17.2 | acute_kidney_injury |
| 35209273 | Acute kidney failure, unspecified | N17.9 | acute_kidney_injury |
| 35209270 | Acute kidney failure with acute cortical necrosis | N17.1 | acute_kidney_injury |
| 35208069 | Acute respiratory distress syndrome | J80 | acute_respiratory_distress_syndrome |
| 35207769 | Left anterior fascicular block | I44.4 | arrythmias |
| 1553751 | Longstanding persistent atrial fibrillation | I48.11 | arrythmias |
| 1569162 | Atrioventricular and left bundle-branch block | I44 | arrythmias |
| 35207790 | Other specified cardiac arrhythmias | I49.8 | arrythmias |
| 45547315 | Encounter for checking and testing of cardiac pacemaker pulse generator [battery] | Z45.010 | arrythmias |
| 1569173 | Atypical atrial flutter | I48.4 | arrythmias |
| 35225413 | Presence of cardiac pacemaker | Z95.0 | arrythmias |
| 35207774 | Trifascicular block | I45.3 | arrythmias |
| 1569170 | Atrial fibrillation and flutter | I48 | arrythmias |
| 35207771 | Left bundle-branch block, unspecified | I44.7 | arrythmias |
| 35207775 | Nonspecific intraventricular block | I45.4 | arrythmias |
| 1569168 | Cardiac arrest | I46 | arrythmias |
| 35207772 | Right fascicular block | I45.0 | arrythmias |
| 35207780 | Re-entry ventricular arrhythmia | I47.0 | arrythmias |
| 35207785 | Persistent atrial fibrillation | I48.1 | arrythmias |
| 1569172 | Typical atrial flutter | I48.3 | arrythmias |
| 35207791 | Cardiac arrhythmia, unspecified | I49.9 | arrythmias |
| 45562352 | Long QT syndrome | I45.81 | arrythmias |
| 35207781 | Supraventricular tachycardia | I47.1 | arrythmias |
| 1569171 | Chronic atrial fibrillation | I48.2 | arrythmias |
| 1569175 | Other cardiac arrhythmias | I49 | arrythmias |
| 1569163 | Other and unspecified atrioventricular block | I44.3 | arrythmias |
| 35207782 | Ventricular tachycardia | I47.2 | arrythmias |
| 35207776 | Other specified heart block | I45.5 | arrythmias |
| 35207777 | Pre-excitation syndrome | I45.6 | arrythmias |
| 45562353 | Ventricular fibrillation | I49.01 | arrythmias |
| 35207786 | Atrial premature depolarization | I49.1 | arrythmias |
| 45576876 | Unspecified atrial fibrillation | I48.91 | arrythmias |
| 35207773 | Bifascicular block | I45.2 | arrythmias |
| 45591468 | Other premature depolarization | I49.49 | arrythmias |
| 35207789 | Sick sinus syndrome | I49.5 | arrythmias |
| 45557544 | Unspecified right bundle-branch block | I45.10 | arrythmias |
| 45581775 | Other fascicular block | I44.69 | arrythmias |
| 1572186 | Abnormalities of heart beat | R00 | arrythmias |
| 45572094 | Unspecified atrial flutter | I48.92 | arrythmias |
| 1569174 | Unspecified atrial fibrillation and atrial flutter | I48.9 | arrythmias |
| 1569177 | Other and unspecified premature depolarization | I49.4 | arrythmias |
| 45605798 | Other specified conduction disorders | I45.89 | arrythmias |
| 1569166 | Other and unspecified right bundle-branch block | I45.1 | arrythmias |
| 45595516 | Encounter for adjustment and management of other part of cardiac pacemaker | Z45.018 | arrythmias |
| 1569165 | Other conduction disorders | I45 | arrythmias |
| 1553753 | Chronic atrial fibrillation, unspecified | I48.20 | arrythmias |
| 35207787 | Junctional premature depolarization | I49.2 | arrythmias |
| 45596205 | Other right bundle-branch block | I45.19 | arrythmias |
| 35207783 | Paroxysmal tachycardia, unspecified | I47.9 | arrythmias |
| 35207768 | Atrioventricular block, complete | I44.2 | arrythmias |
| 45562351 | Unspecified fascicular block | I44.60 | arrythmias |
| 35211263 | Palpitations | R00.2 | arrythmias |
| 35207778 | Conduction disorder, unspecified | I45.9 | arrythmias |
| 35211264 | Other abnormalities of heart beat | R00.8 | arrythmias |
| 1569176 | Ventricular fibrillation and flutter | I49.0 | arrythmias |
| 45552792 | Other atrioventricular block | I44.39 | arrythmias |
| 35207766 | Atrioventricular block, first degree | I44.0 | arrythmias |
| 35207767 | Atrioventricular block, second degree | I44.1 | arrythmias |
| 1569169 | Paroxysmal tachycardia | I47 | arrythmias |
| 35211261 | Tachycardia, unspecified | R00.0 | arrythmias |
| 35207770 | Left posterior fascicular block | I44.5 | arrythmias |
| 1553754 | Permanent atrial fibrillation | I48.21 | arrythmias |
| 35211262 | Bradycardia, unspecified | R00.1 | arrythmias |
| 1569167 | Other specified conduction disorders | I45.8 | arrythmias |
| 45577776 | Unspecified abnormalities of heart beat | R00.9 | arrythmias |
| 45552796 | Unspecified premature depolarization | I49.40 | arrythmias |
| 45596204 | Unspecified atrioventricular block | I44.30 | arrythmias |
| 1553752 | Other persistent atrial fibrillation | I48.19 | arrythmias |
| 45533455 | Ventricular flutter | I49.02 | arrythmias |
| 1569164 | Other and unspecified fascicular block | I44.6 | arrythmias |
| 35207784 | Paroxysmal atrial fibrillation | I48.0 | arrythmias |
| 35207788 | Ventricular premature depolarization | I49.3 | arrythmias |
| 35211265 | Benign and innocent cardiac murmurs | R01.0 | cardiovascular_signs_and_sx |
| 1572187 | Cardiac murmurs and other cardiac sounds | R01 | cardiovascular_signs_and_sx |
| 35211267 | Other cardiac sounds | R01.2 | cardiovascular_signs_and_sx |
| 35211268 | Elevated blood-pressure reading, without diagnosis of hypertension | R03.0 | cardiovascular_signs_and_sx |
| 35211269 | Nonspecific low blood-pressure reading | R03.1 | cardiovascular_signs_and_sx |
| 45558447 | Acute idiopathic pulmonary hemorrhage in infants | R04.81 | cardiovascular_signs_and_sx |
| 35211266 | Cardiac murmur, unspecified | R01.1 | cardiovascular_signs_and_sx |
| 1572188 | Abnormal blood-pressure reading, without diagnosis | R03 | cardiovascular_signs_and_sx |
| 35211314 | Cyanosis | R23.0 | cardiovascular_signs_and_sx |
| 35211352 | Parosmia | R43.1 | changes_in_taste_and_smell |
| 35211353 | Parageusia | R43.2 | changes_in_taste_and_smell |
| 45573025 | Unspecified disturbances of smell and taste | R43.9 | changes_in_taste_and_smell |
| 35211351 | Anosmia | R43.0 | changes_in_taste_and_smell |
| 35211354 | Other disturbances of smell and taste | R43.8 | changes_in_taste_and_smell |
| 1572242 | Disturbances of smell and taste | R43 | changes_in_taste_and_smell |
| 45534424 | Chest pain, unspecified | R07.9 | chest_pain |
| 35211284 | Chest pain on breathing | R07.1 | chest_pain |
| 45597167 | Intercostal pain | R07.82 | chest_pain |
| 45602002 | Other chest pain | R07.89 | chest_pain |
| 1572194 | Other chest pain | R07.8 | chest_pain |
| 35211283 | Pain in throat | R07.0 | chest_pain |
| 45587497 | Pleurodynia | R07.81 | chest_pain |
| 35211285 | Precordial pain | R07.2 | chest_pain |
| 1572193 | Pain in throat and chest | R07 | chest_pain |
| 35207245 | Expressive language disorder | F80.1 | cognitive_function |
| 1572239 | Other symptoms and signs involving cognitive functions and awareness | R41 | cognitive_function |
| 35211349 | Other amnesia | R41.3 | cognitive_function |
| 35225170 | Problems related to education and literacy, unspecified | Z55.9 | cognitive_function |
| 45587018 | Muscle weakness (generalized) | M62.81 | fatigue_and_malaise |
| 45582718 | Other malaise | R53.81 | fatigue_and_malaise |
| 45534458 | Other fatigue | R53.83 | fatigue_and_malaise |
| 35207505 | Postviral fatigue syndrome | G93.3 | fatigue_and_malaise |
| 1572256 | Other malaise and fatigue | R53.8 | fatigue_and_malaise |
| 45573032 | Chronic fatigue, unspecified | R53.82 | fatigue_and_malaise |
| 45542780 | Transfusion associated circulatory overload | E87.71 | fluid_and_electrolyte |
| 35207094 | Acidosis | E87.2 | fluid_and_electrolyte |
| 45576480 | Fluid overload, unspecified | E87.70 | fluid_and_electrolyte |
| 35207099 | Fluid overload | E87.7 | fluid_and_electrolyte |
| 1568078 | Other disorders of fluid, electrolyte and acid-base balance | E87 | fluid_and_electrolyte |
| 35207100 | Other disorders of electrolyte and fluid balance, not elsewhere classified | E87.8 | fluid_and_electrolyte |
| 35207093 | Hypo-osmolality and hyponatremia | E87.1 | fluid_and_electrolyte |
| 35207095 | Alkalosis | E87.3 | fluid_and_electrolyte |
| 35207096 | Mixed disorder of acid-base balance | E87.4 | fluid_and_electrolyte |
| 35207098 | Hypokalemia | E87.6 | fluid_and_electrolyte |
| 45542781 | Other fluid overload | E87.79 | fluid_and_electrolyte |
| 35207097 | Hyperkalemia | E87.5 | fluid_and_electrolyte |
| 35207092 | Hyperosmolality and hypernatremia | E87.0 | fluid_and_electrolyte |
| 45562109 | Chronic pain syndrome | G89.4 | generalized_pain |
| 35211389 | Pain, unspecified | R52 | generalized_pain |
| 1568420 | Pain, not elsewhere classified | G89 | generalized_pain |
| 45566906 | Central pain syndrome | G89.0 | generalized_pain |
| 45542912 | Other chronic pain | G89.29 | generalized_pain |
| 45547413 | Syphilitic alopecia | A51.32 | hair_loss |
| 1569792 | Alopecia areata | L63 | hair_loss |
| 35208591 | Alopecia (capitis) totalis | L63.0 | hair_loss |
| 35208592 | Alopecia universalis | L63.1 | hair_loss |
| 35208593 | Ophiasis | L63.2 | hair_loss |
| 35208594 | Other alopecia areata | L63.8 | hair_loss |
| 35208595 | Alopecia areata, unspecified | L63.9 | hair_loss |
| 1569793 | Androgenic alopecia | L64 | hair_loss |
| 35208596 | Drug-induced androgenic alopecia | L64.0 | hair_loss |
| 35208597 | Other androgenic alopecia | L64.8 | hair_loss |
| 35208598 | Androgenic alopecia, unspecified | L64.9 | hair_loss |
| 1569794 | Other nonscarring hair loss | L65 | hair_loss |
| 35208599 | Telogen effluvium | L65.0 | hair_loss |
| 35208600 | Anagen effluvium | L65.1 | hair_loss |
| 35208601 | Alopecia mucinosa | L65.2 | hair_loss |
| 35208602 | Other specified nonscarring hair loss | L65.8 | hair_loss |
| 35208603 | Nonscarring hair loss, unspecified | L65.9 | hair_loss |
| 1569795 | Cicatricial alopecia [scarring hair loss] | L66 | hair_loss |
| 35208604 | Pseudopelade | L66.0 | hair_loss |
| 35208605 | Lichen planopilaris | L66.1 | hair_loss |
| 35208606 | Folliculitis decalvans | L66.2 | hair_loss |
| 35208607 | Perifolliculitis capitis abscedens | L66.3 | hair_loss |
| 35208608 | Folliculitis ulerythematosa reticulata | L66.4 | hair_loss |
| 35208609 | Other cicatricial alopecia | L66.8 | hair_loss |
| 35208610 | Cicatricial alopecia, unspecified | L66.9 | hair_loss |
| 45566877 | Ophthalmoplegic migraine, intractable, with status migrainosus | G43.B11 | headache |
| 45562091 | Chronic tension-type headache | G44.22 | headache |
| 45562088 | Hemiplegic migraine | G43.4 | headache |
| 1568355 | Post-traumatic headache, unspecified | G44.30 | headache |
| 35207382 | Status migrainosus | G43.2 | headache |
| 45547740 | Drug-induced headache, not elsewhere classified, intractable | G44.41 | headache |
| 45562090 | Periodic headache syndromes in child or adult, intractable, with status migrainosus | G43.C11 | headache |
| 45591179 | Cyclical vomiting, not intractable, with status migrainosus | G43.A01 | headache |
| 45547737 | Chronic paroxysmal hemicrania, not intractable | G44.049 | headache |
| 1568341 | Other migraine, intractable | G43.81 | headache |
| 1568342 | Menstrual migraine, not intractable | G43.82 | headache |
| 35207389 | Other migraine, intractable | G43.891 | headache |
| 1568352 | Tension-type headache | G44.2 | headache |
| 725459 | Headache, unspecified | R51.9 | headache |
| 35207384 | Hemiplegic migraine, intractable | G43.41 | headache |
| 45533136 | Migraine without aura, not intractable, without status migrainosus | G43.009 | headache |
| 35207385 | Persistent migraine aura without cerebral infarction, not intractable | G43.50 | headache |
| 1568344 | Migraine, unspecified | G43.9 | headache |
| 35207390 | Other migraine, not intractable | G43.899 | headache |
| 45586294 | Primary thunderclap headache | G44.53 | headache |
| 35207381 | Migraine with aura, intractable | G43.11 | headache |
| 45538119 | Drug-induced headache, not elsewhere classified, not intractable | G44.40 | headache |
| 1568339 | Other migraine | G43.8 | headache |
| 35207380 | Migraine with aura, not intractable | G43.10 | headache |
| 45576582 | Post-traumatic headache, unspecified, intractable | G44.301 | headache |
| 45605550 | Chronic cluster headache, intractable | G44.021 | headache |
| 1568335 | Persistent migraine aura with cerebral infarction, not intractable | G43.60 | headache |
| 45566881 | Episodic tension-type headache, not intractable | G44.219 | headache |
| 45566879 | Chronic cluster headache | G44.02 | headache |
| 45542890 | Chronic tension-type headache, not intractable | G44.229 | headache |
| 35207391 | Migraine, unspecified, not intractable | G43.90 | headache |
| 45552531 | Vascular headache, not elsewhere classified, not intractable | G44.10 | headache |
| 45566876 | Ophthalmoplegic migraine | G43.B | headache |
| 45591178 | Chronic migraine without aura, not intractable, without status migrainosus | G43.709 | headache |
| 1568357 | Drug-induced headache, not elsewhere classified | G44.4 | headache |
| 45562089 | Cyclical vomiting, intractable, with status migrainosus | G43.A11 | headache |
| 45576585 | Primary stabbing headache | G44.85 | headache |
| 1568358 | Complicated headache syndromes | G44.5 | headache |
| 1568354 | Post-traumatic headache | G44.3 | headache |
| 45591181 | Periodic headache syndromes in child or adult, not intractable | G43.C0 | headache |
| 35211388 | Headache | R51 | headache |
| 1568347 | Cluster headache syndrome, unspecified | G44.00 | headache |
| 766354 | Cervicogenic headache | G44.86 | headache |
| 45538112 | Cyclical vomiting, in migraine, not intractable | G43.A0 | headache |
| 45600790 | Primary exertional headache | G44.84 | headache |
| 45542887 | Migraine, unspecified, not intractable, with status migrainosus | G43.901 | headache |
| 45552528 | Other migraine, not intractable, with status migrainosus | G43.801 | headache |
| 45533138 | Chronic migraine without aura, not intractable, with status migrainosus | G43.701 | headache |
| 1568337 | Chronic migraine without aura | G43.7 | headache |
| 45595944 | Menstrual migraine, not intractable, with status migrainosus | G43.D01 | headache |
| 45591180 | Ophthalmoplegic migraine, intractable, without status migrainosus | G43.B19 | headache |
| 1568349 | Short lasting unilateral neuralgiform headache with conjunctival injection and tearing (SUNCT) | G44.05 | headache |
| 1568346 | Cluster headaches and other trigeminal autonomic cephalgias (TAC) | G44.0 | headache |
| 45542891 | Chronic post-traumatic headache, intractable | G44.321 | headache |
| 1568359 | Other specified headache syndromes | G44.8 | headache |
| 45605545 | Persistent migraine aura with cerebral infarction, not intractable, without status migrainosus | G43.609 | headache |
| 45547732 | Hemiplegic migraine, not intractable, with status migrainosus | G43.401 | headache |
| 45557237 | Other migraine, intractable, with status migrainosus | G43.811 | headache |
| 45581494 | Periodic headache syndromes in child or adult, not intractable, with status migrainosus | G43.C01 | headache |
| 45576580 | Chronic paroxysmal hemicrania | G44.04 | headache |
| 45571807 | Chronic cluster headache, not intractable | G44.029 | headache |
| 45538110 | Hemiplegic migraine, intractable, without status migrainosus | G43.419 | headache |
| 45586292 | Other migraine, not intractable, without status migrainosus | G43.809 | headache |
| 45557236 | Migraine without aura, intractable, without status migrainosus | G43.019 | headache |
| 45533142 | Episodic cluster headache, not intractable | G44.019 | headache |
| 35207379 | Migraine without aura, intractable | G43.01 | headache |
| 45576578 | Menstrual migraine, intractable, with status migrainosus | G43.D11 | headache |
| 45552529 | Ophthalmoplegic migraine, not intractable, with status migrainosus | G43.B01 | headache |
| 45547735 | Ophthalmoplegic migraine, not intractable | G43.B0 | headache |
| 1568356 | Acute post-traumatic headache | G44.31 | headache |
| 45591182 | Menstrual migraine, intractable, without status migrainosus | G43.D19 | headache |
| 35207383 | Hemiplegic migraine, not intractable | G43.40 | headache |
| 35207388 | Chronic migraine without aura, intractable | G43.71 | headache |
| 45542888 | Short lasting unilateral neuralgiform headache with conjunctival injection and tearing (SUNCT), intractable | G44.051 | headache |
| 45605543 | Migraine with aura, not intractable, with status migrainosus | G43.101 | headache |
| 45566880 | Episodic tension-type headache, intractable | G44.211 | headache |
| 35207386 | Persistent migraine aura without cerebral infarction, intractable | G43.51 | headache |
| 1568353 | Tension-type headache, unspecified | G44.20 | headache |
| 45562087 | Migraine with aura, not intractable, without status migrainosus | G43.109 | headache |
| 45552533 | Hemicrania continua | G44.51 | headache |
| 45538115 | Periodic headache syndromes in child or adult, intractable | G43.C1 | headache |
| 45605551 | Chronic paroxysmal hemicrania, intractable | G44.041 | headache |
| 45552532 | Chronic post-traumatic headache | G44.32 | headache |
| 45586289 | Hemiplegic migraine, intractable, with status migrainosus | G43.411 | headache |
| 45605547 | Cyclical vomiting, in migraine, intractable | G43.A1 | headache |
| 45538117 | Acute post-traumatic headache, intractable | G44.311 | headache |
| 45600786 | Migraine, unspecified, not intractable, without status migrainosus | G43.909 | headache |
| 45566875 | Migraine with aura, intractable, with status migrainosus | G43.111 | headache |
| 45547739 | Episodic tension-type headache | G44.21 | headache |
| 45571805 | Migraine, unspecified, intractable, without status migrainosus | G43.919 | headache |
| 45571804 | Menstrual migraine, intractable, with status migrainosus | G43.831 | headache |
| 45547734 | Persistent migraine aura with cerebral infarction, intractable, without status migrainosus | G43.619 | headache |
| 45576584 | Primary cough headache | G44.83 | headache |
| 45581491 | Chronic migraine without aura, intractable, with status migrainosus | G43.711 | headache |
| 45538118 | Chronic post-traumatic headache, not intractable | G44.329 | headache |
| 45600788 | Episodic paroxysmal hemicrania, not intractable | G44.039 | headache |
| 45533139 | Cyclical vomiting, not intractable, without status migrainosus | G43.A09 | headache |
| 45586291 | Persistent migraine aura without cerebral infarction, intractable, without status migrainosus | G43.519 | headache |
| 1568336 | Persistent migraine aura with cerebral infarction, intractable | G43.61 | headache |
| 45538108 | Migraine without aura, intractable, with status migrainosus | G43.011 | headache |
| 45576577 | Periodic headache syndromes in child or adult, intractable, without status migrainosus | G43.C19 | headache |
| 45566882 | Post-traumatic headache, unspecified, not intractable | G44.309 | headache |
| 45571802 | Menstrual migraine, not intractable, with status migrainosus | G43.821 | headache |
| 1568340 | Other migraine, not intractable | G43.80 | headache |
| 45586293 | Episodic cluster headache | G44.01 | headache |
| 35207378 | Migraine without aura, not intractable | G43.00 | headache |
| 45591184 | Hypnic headache | G44.81 | headache |
| 1568331 | Migraine without aura | G43.0 | headache |
| 45600789 | Other complicated headache syndrome | G44.59 | headache |
| 45547738 | Tension-type headache, unspecified, not intractable | G44.209 | headache |
| 1568332 | Migraine with aura | G43.1 | headache |
| 45605546 | Persistent migraine aura with cerebral infarction, intractable, with status migrainosus | G43.611 | headache |
| 45576581 | Other trigeminal autonomic cephalgias (TAC), not intractable | G44.099 | headache |
| 725458 | Headache with orthostatic component, not elsewhere classified | R51.0 | headache |
| 45605549 | Episodic cluster headache, intractable | G44.011 | headache |
| 45605553 | Chronic tension-type headache, intractable | G44.221 | headache |
| 1568350 | Other trigeminal autonomic cephalgias (TAC) | G44.09 | headache |
| 45566878 | Cluster headache syndrome, unspecified, intractable | G44.001 | headache |
| 45538116 | Menstrual migraine, not intractable, without status migrainosus | G43.D09 | headache |
| 1568345 | Other headache syndromes | G44 | headache |
| 45557238 | Migraine, unspecified, intractable, with status migrainosus | G43.911 | headache |
| 1568351 | Vascular headache, not elsewhere classified | G44.1 | headache |
| 45576583 | New daily persistent headache (NDPH) | G44.52 | headache |
| 45538109 | Hemiplegic migraine, not intractable, without status migrainosus | G43.409 | headache |
| 45552527 | Migraine with aura, intractable, without status migrainosus | G43.119 | headache |
| 45533137 | Persistent migraine aura without cerebral infarction, not intractable, without status migrainosus | G43.509 | headache |
| 45566874 | Migraine without aura, not intractable, with status migrainosus | G43.001 | headache |
| 1568348 | Episodic paroxysmal hemicrania | G44.03 | headache |
| 45538114 | Ophthalmoplegic migraine, intractable | G43.B1 | headache |
| 45605552 | Short lasting unilateral neuralgiform headache with conjunctival injection and tearing (SUNCT), not intractable | G44.059 | headache |
| 1568333 | Persistent migraine aura without cerebral infarction | G43.5 | headache |
| 45571803 | Menstrual migraine, not intractable, without status migrainosus | G43.829 | headache |
| 45595945 | Vascular headache, not elsewhere classified, intractable | G44.11 | headache |
| 45571806 | Periodic headache syndromes in child or adult, not intractable, without status migrainosus | G43.C09 | headache |
| 1568343 | Menstrual migraine, intractable | G43.83 | headache |
| 1568334 | Persistent migraine aura with cerebral infarction | G43.6 | headache |
| 35207392 | Migraine, unspecified, intractable | G43.91 | headache |
| 45605548 | Cluster headache syndrome, unspecified, not intractable | G44.009 | headache |
| 45600785 | Chronic migraine without aura, intractable, without status migrainosus | G43.719 | headache |
| 45542889 | Tension-type headache, unspecified, intractable | G44.201 | headache |
| 35207387 | Chronic migraine without aura, not intractable | G43.70 | headache |
| 45547733 | Persistent migraine aura with cerebral infarction, not intractable, with status migrainosus | G43.601 | headache |
| 45586290 | Persistent migraine aura without cerebral infarction, not intractable, with status migrainosus | G43.501 | headache |
| 45533143 | Other trigeminal autonomic cephalgias (TAC), intractable | G44.091 | headache |
| 1568330 | Migraine | G43 | headache |
| 45605554 | Acute post-traumatic headache, not intractable | G44.319 | headache |
| 45595940 | Persistent migraine aura without cerebral infarction, intractable, with status migrainosus | G43.511 | headache |
| 45591183 | Episodic paroxysmal hemicrania, intractable | G44.031 | headache |
| 45595942 | Menstrual migraine, intractable, without status migrainosus | G43.839 | headache |
| 45538111 | Other migraine, intractable, without status migrainosus | G43.819 | headache |
| 1568338 | Periodic headache syndromes in child or adult | G43.C | headache |
| 45533144 | Other headache syndrome | G44.89 | headache |
| 45538113 | Ophthalmoplegic migraine, not intractable, without status migrainosus | G43.B09 | headache |
| 1569168 | Cardiac arrest | I46 | heart_disease |
| 35207800 | Cardiomegaly | I51.7 | heart_disease |
| 45576878 | Chronic systolic (congestive) heart failure | I50.22 | heart_disease |
| 35207796 | Rupture of papillary muscle, not elsewhere classified | I51.2 | heart_disease |
| 35207740 | Nonrheumatic tricuspid (valve) stenosis with insufficiency | I36.2 | heart_disease |
| 1326605 | Right heart failure due to left heart failure | I50.814 | heart_disease |
| 45586587 | Unspecified systolic (congestive) heart failure | I50.20 | heart_disease |
| 35207718 | Acute pericarditis, unspecified | I30.9 | heart_disease |
| 1326606 | Biventricular heart failure | I50.82 | heart_disease |
| 35207724 | Disease of pericardium, unspecified | I31.9 | heart_disease |
| 1326608 | End stage heart failure | I50.84 | heart_disease |
| 35207739 | Nonrheumatic tricuspid (valve) insufficiency | I36.1 | heart_disease |
| 45538387 | Takotsubo syndrome | I51.81 | heart_disease |
| 35207730 | Nonrheumatic mitral (valve) stenosis | I34.2 | heart_disease |
| 45543182 | Acute systolic (congestive) heart failure | I50.21 | heart_disease |
| 1326601 | Right heart failure, unspecified | I50.810 | heart_disease |
| 1326607 | High output heart failure | I50.83 | heart_disease |
| 35207731 | Other nonrheumatic mitral valve disorders | I34.8 | heart_disease |
| 35207732 | Nonrheumatic mitral valve disorder, unspecified | I34.9 | heart_disease |
| 45533457 | Unspecified combined systolic (congestive) and diastolic (congestive) heart failure | I50.40 | heart_disease |
| 1569154 | Other diseases of pericardium | I31 | heart_disease |
| 45591469 | Acute combined systolic (congestive) and diastolic (congestive) heart failure | I50.41 | heart_disease |
| 35207755 | Dilated cardiomyopathy | I42.0 | heart_disease |
| 35207799 | Myocardial degeneration | I51.5 | heart_disease |
| 35207792 | Left ventricular failure, unspecified | I50.1 | heart_disease |
| 35207756 | Obstructive hypertrophic cardiomyopathy | I42.1 | heart_disease |
| 45601038 | Unspecified diastolic (congestive) heart failure | I50.30 | heart_disease |
| 45567183 | Other ill-defined heart diseases | I51.89 | heart_disease |
| 45567181 | Acute on chronic combined systolic (congestive) and diastolic (congestive) heart failure | I50.43 | heart_disease |
| 35207720 | Chronic constrictive pericarditis | I31.1 | heart_disease |
| 35207738 | Nonrheumatic tricuspid (valve) stenosis | I36.0 | heart_disease |
| 45567180 | Acute on chronic systolic (congestive) heart failure | I50.23 | heart_disease |
| 35207798 | Myocarditis, unspecified | I51.4 | heart_disease |
| 35207729 | Nonrheumatic mitral (valve) prolapse | I34.1 | heart_disease |
| 35207723 | Other specified diseases of pericardium | I31.8 | heart_disease |
| 45562355 | Acute on chronic diastolic (congestive) heart failure | I50.33 | heart_disease |
| 1326604 | Acute on chronic right heart failure | I50.813 | heart_disease |
| 45548022 | Acute diastolic (congestive) heart failure | I50.31 | heart_disease |
| 1569178 | Heart failure | I50 | heart_disease |
| 35207795 | Rupture of chordae tendineae, not elsewhere classified | I51.1 | heart_disease |
| 35207719 | Chronic adhesive pericarditis | I31.0 | heart_disease |
| 35207742 | Nonrheumatic tricuspid valve disorder, unspecified | I36.9 | heart_disease |
| 45586588 | Chronic combined systolic (congestive) and diastolic (congestive) heart failure | I50.42 | heart_disease |
| 35207802 | Other heart disorders in diseases classified elsewhere | I52 | heart_disease |
| 35207757 | Other hypertrophic cardiomyopathy | I42.2 | heart_disease |
| 1326609 | Other heart failure | I50.89 | heart_disease |
| 1569156 | Nonrheumatic mitral valve disorders | I34 | heart_disease |
| 1569182 | Complications and ill-defined descriptions of heart disease | I51 | heart_disease |
| 35207722 | Pericardial effusion (noninflammatory) | I31.3 | heart_disease |
| 35207763 | Other cardiomyopathies | I42.8 | heart_disease |
| 35207801 | Heart disease, unspecified | I51.9 | heart_disease |
| 35207741 | Other nonrheumatic tricuspid valve disorders | I36.8 | heart_disease |
| 35207793 | Heart failure, unspecified | I50.9 | heart_disease |
| 35207794 | Cardiac septal defect, acquired | I51.0 | heart_disease |
| 35207728 | Nonrheumatic mitral (valve) insufficiency | I34.0 | heart_disease |
| 1569161 | Cardiomyopathy | I42 | heart_disease |
| 45557538 | Coronary artery aneurysm | I25.41 | heart_disease |
| 35207764 | Cardiomyopathy, unspecified | I42.9 | heart_disease |
| 35207758 | Endomyocardial (eosinophilic) disease | I42.3 | heart_disease |
| 35207759 | Endocardial fibroelastosis | I42.4 | heart_disease |
| 1326603 | Chronic right heart failure | I50.812 | heart_disease |
| 35207760 | Other restrictive cardiomyopathy | I42.5 | heart_disease |
| 45533456 | Chronic diastolic (congestive) heart failure | I50.32 | heart_disease |
| 1569158 | Nonrheumatic tricuspid valve disorders | I36 | heart_disease |
| 1326602 | Acute right heart failure | I50.811 | heart_disease |
| 35207636 | Acute rheumatic pericarditis | I01.0 | heart_disease |
| 45571465 | Viral pericarditis | B33.23 | heart_disease |
| 1569153 | Acute pericarditis | I30 | heart_disease |
| 35207715 | Acute nonspecific idiopathic pericarditis | I30.0 | heart_disease |
| 35207716 | Infective pericarditis | I30.1 | heart_disease |
| 35207717 | Other forms of acute pericarditis | I30.8 | heart_disease |
| 35207725 | Pericarditis in diseases classified elsewhere | I32 | heart_disease |
| 35207666 | Chronic rheumatic pericarditis | I09.2 | heart_disease |
| 35207919 | Postcardiotomy syndrome | I97.0 | heart_disease |
| 45572430 | Pain in left hip | M25.552 | musculoskeletal |
| 45577229 | Stiffness of left shoulder, not elsewhere classified | M25.612 | musculoskeletal |
| 45548444 | Occipital neuralgia | M54.81 | musculoskeletal |
| 45553157 | Pain in left elbow | M25.522 | musculoskeletal |
| 45577230 | Stiffness of right elbow, not elsewhere classified | M25.621 | musculoskeletal |
| 1571058 | Pain in limb, hand, foot, fingers and toes | M79.6 | musculoskeletal |
| 1570567 | Pain in hip | M25.55 | musculoskeletal |
| 1570568 | Pain in knee | M25.56 | musculoskeletal |
| 45557903 | Other dorsalgia | M54.89 | musculoskeletal |
| 1595620 | Myalgia, other site | M79.18 | musculoskeletal |
| 45582249 | Pain in right hand | M79.641 | musculoskeletal |
| 45572432 | Stiffness of right wrist, not elsewhere classified | M25.631 | musculoskeletal |
| 45533940 | Pain in left forearm | M79.632 | musculoskeletal |
| 45606202 | Pain in wrist | M25.53 | musculoskeletal |
| 725390 | Stiffness of other specified joint, not elsewhere classified | M25.69 | musculoskeletal |
| 45591807 | Pain in right shoulder | M25.511 | musculoskeletal |
| 45577227 | Pain in unspecified joint | M25.50 | musculoskeletal |
| 766414 | Other low back pain | M54.59 | musculoskeletal |
| 45538866 | Pain in unspecified hand | M79.643 | musculoskeletal |
| 45557858 | Stiffness of unspecified ankle, not elsewhere classified | M25.673 | musculoskeletal |
| 45567529 | Stiffness of right shoulder, not elsewhere classified | M25.611 | musculoskeletal |
| 35208969 | Low back pain | M54.5 | musculoskeletal |
| 45587066 | Pain in right lower leg | M79.661 | musculoskeletal |
| 45587065 | Pain in left thigh | M79.652 | musculoskeletal |
| 1595617 | Myalgia, unspecified site | M79.10 | musculoskeletal |
| 37200700 | Pain in joints of hand | M25.54 | musculoskeletal |
| 45582116 | Pain in left ankle and joints of left foot | M25.572 | musculoskeletal |
| 45596549 | Pain in right knee | M25.561 | musculoskeletal |
| 45567533 | Stiffness of left foot, not elsewhere classified | M25.675 | musculoskeletal |
| 37200701 | Pain in joints of right hand | M25.541 | musculoskeletal |
| 45582247 | Pain in left leg | M79.605 | musculoskeletal |
| 45562838 | Pain in unspecified limb | M79.609 | musculoskeletal |
| 45543690 | Pain in right forearm | M79.631 | musculoskeletal |
| 45591935 | Pain in right thigh | M79.651 | musculoskeletal |
| 45572434 | Stiffness of left ankle, not elsewhere classified | M25.672 | musculoskeletal |
| 766412 | Low back pain, unspecified | M54.50 | musculoskeletal |
| 45586943 | Stiffness of left wrist, not elsewhere classified | M25.632 | musculoskeletal |
| 45538740 | Pain in right ankle and joints of right foot | M25.571 | musculoskeletal |
| 45562839 | Pain in right upper arm | M79.621 | musculoskeletal |
| 45567532 | Stiffness of right ankle, not elsewhere classified | M25.671 | musculoskeletal |
| 45538867 | Pain in right finger(s) | M79.644 | musculoskeletal |
| 45557993 | Pain in unspecified foot | M79.673 | musculoskeletal |
| 766413 | Vertebrogenic low back pain | M54.51 | musculoskeletal |
| 45553161 | Stiffness of unspecified hand, not elsewhere classified | M25.649 | musculoskeletal |
| 45533939 | Pain in right leg | M79.604 | musculoskeletal |
| 45601546 | Pain in right toe(s) | M79.674 | musculoskeletal |
| 45587067 | Pain in unspecified lower leg | M79.669 | musculoskeletal |
| 45591934 | Pain in unspecified finger(s) | M79.646 | musculoskeletal |
| 45557992 | Pain in right arm | M79.601 | musculoskeletal |
| 35209013 | Soft tissue disorder, unspecified | M79.9 | musculoskeletal |
| 45572433 | Stiffness of unspecified hip, not elsewhere classified | M25.659 | musculoskeletal |
| 45572552 | Pain in left lower leg | M79.662 | musculoskeletal |
| 35208970 | Pain in thoracic spine | M54.6 | musculoskeletal |
| 45553159 | Stiffness of unspecified elbow, not elsewhere classified | M25.629 | musculoskeletal |
| 45587064 | Pain in arm, unspecified | M79.603 | musculoskeletal |
| 45572431 | Pain in unspecified ankle and joints of unspecified foot | M25.579 | musculoskeletal |
| 45586944 | Stiffness of unspecified wrist, not elsewhere classified | M25.639 | musculoskeletal |
| 45596550 | Stiffness of left hand, not elsewhere classified | M25.642 | musculoskeletal |
| 45557857 | Stiffness of right knee, not elsewhere classified | M25.661 | musculoskeletal |
| 45606201 | Pain in unspecified elbow | M25.529 | musculoskeletal |
| 45577231 | Stiffness of left knee, not elsewhere classified | M25.662 | musculoskeletal |
| 45553271 | Pain in unspecified upper arm | M79.629 | musculoskeletal |
| 45582118 | Stiffness of unspecified foot, not elsewhere classified | M25.676 | musculoskeletal |
| 45606200 | Pain in unspecified shoulder | M25.519 | musculoskeletal |
| 37200703 | Pain in joints of unspecified hand | M25.549 | musculoskeletal |
| 45553158 | Pain in left wrist | M25.532 | musculoskeletal |
| 1570569 | Pain in ankle and joints of foot | M25.57 | musculoskeletal |
| 45562840 | Pain in right foot | M79.671 | musculoskeletal |
| 45567531 | Stiffness of unspecified knee, not elsewhere classified | M25.669 | musculoskeletal |
| 45606323 | Pain in left finger(s) | M79.645 | musculoskeletal |
| 45591810 | Stiffness of left hip, not elsewhere classified | M25.652 | musculoskeletal |
| 45591936 | Pain in left toe(s) | M79.675 | musculoskeletal |
| 45562837 | Pain in left arm | M79.602 | musculoskeletal |
| 45591809 | Pain in elbow | M25.52 | musculoskeletal |
| 35208971 | Dorsalgia, unspecified | M54.9 | musculoskeletal |
| 35209006 | Myalgia | M79.1 | musculoskeletal |
| 45557856 | Pain in unspecified wrist | M25.539 | musculoskeletal |
| 45577228 | Pain in unspecified hip | M25.559 | musculoskeletal |
| 1570565 | Pain in joint | M25.5 | musculoskeletal |
| 45553160 | Stiffness of right hand, not elsewhere classified | M25.641 | musculoskeletal |
| 45601545 | Pain in unspecified thigh | M79.659 | musculoskeletal |
| 45596551 | Stiffness of right foot, not elsewhere classified | M25.674 | musculoskeletal |
| 45533816 | Pain in left knee | M25.562 | musculoskeletal |
| 45586942 | Stiffness of unspecified joint, not elsewhere classified | M25.60 | musculoskeletal |
| 45577358 | Pain in unspecified forearm | M79.639 | musculoskeletal |
| 45543689 | Pain in leg, unspecified | M79.606 | musculoskeletal |
| 37200702 | Pain in joints of left hand | M25.542 | musculoskeletal |
| 45533941 | Pain in unspecified toe(s) | M79.676 | musculoskeletal |
| 45601424 | Stiffness of right hip, not elsewhere classified | M25.651 | musculoskeletal |
| 1595618 | Myalgia of mastication muscle | M79.11 | musculoskeletal |
| 45548395 | Pain in unspecified knee | M25.569 | musculoskeletal |
| 45557855 | Pain in right wrist | M25.531 | musculoskeletal |
| 45562699 | Pain in right hip | M25.551 | musculoskeletal |
| 45582117 | Stiffness of unspecified shoulder, not elsewhere classified | M25.619 | musculoskeletal |
| 45538868 | Pain in left foot | M79.672 | musculoskeletal |
| 45567530 | Stiffness of left elbow, not elsewhere classified | M25.622 | musculoskeletal |
| 45582248 | Pain in left upper arm | M79.622 | musculoskeletal |
| 45601423 | Pain in right elbow | M25.521 | musculoskeletal |
| 45591808 | Pain in left shoulder | M25.512 | musculoskeletal |
| 725389 | Pain in other specified joint | M25.59 | musculoskeletal |
| 45582250 | Pain in left hand | M79.642 | musculoskeletal |
| 1595619 | Myalgia of auxiliary muscles, head and neck | M79.12 | musculoskeletal |
| 1570566 | Pain in shoulder | M25.51 | musculoskeletal |
| 35207753 | Acute myocarditis, unspecified | I40.9 | myocarditis |
| 45581156 | Viral myocarditis | B33.22 | myocarditis |
| 35207752 | Other acute myocarditis | I40.8 | myocarditis |
| 35207750 | Infective myocarditis | I40.0 | myocarditis |
| 35207754 | Myocarditis in diseases classified elsewhere | I41 | myocarditis |
| 45548447 | Infective myositis, unspecified foot | M60.075 | myositis |
| 1570722 | Interstitial myositis, thigh | M60.15 | myositis |
| 45562755 | Other myositis, left hand | M60.842 | myositis |
| 1570708 | Infective myositis | M60.0 | myositis |
| 45557906 | Infective myositis, right lower leg | M60.061 | myositis |
| 45553208 | Infective myositis, right ankle | M60.070 | myositis |
| 45606255 | Other myositis, unspecified lower leg | M60.869 | myositis |
| 45567581 | Infective myositis, unspecified arm | M60.002 | myositis |
| 45538796 | Infective myositis, other site | M60.08 | myositis |
| 45557904 | Infective myositis, unspecified left leg | M60.004 | myositis |
| 45548402 | Polymyositis, organ involvement unspecified | M33.20 | myositis |
| 45557905 | Infective myositis, left forearm | M60.032 | myositis |
| 45606250 | Infective myositis, unspecified forearm | M60.039 | myositis |
| 45577294 | Other myositis, unspecified forearm | M60.839 | myositis |
| 1570718 | Interstitial myositis, shoulder | M60.11 | myositis |
| 1570709 | Infective myositis, unspecified site | M60.00 | myositis |
| 45557913 | Other myositis, unspecified ankle and foot | M60.879 | myositis |
| 45557908 | Interstitial myositis, unspecified thigh | M60.159 | myositis |
| 1570738 | Other myositis, thigh | M60.85 | myositis |
| 45606248 | Infective myositis, unspecified right leg | M60.003 | myositis |
| 45567582 | Infective myositis, unspecified site | M60.009 | myositis |
| 45596603 | Interstitial myositis, left forearm | M60.132 | myositis |
| 45606251 | Interstitial myositis, right lower leg | M60.161 | myositis |
| 1570720 | Interstitial myositis, forearm | M60.13 | myositis |
| 45567588 | Other myositis, right lower leg | M60.861 | myositis |
| 45596606 | Interstitial myositis, left hand | M60.142 | myositis |
| 1570712 | Infective myositis, forearm | M60.03 | myositis |
| 45577293 | Interstitial myositis, right ankle and foot | M60.171 | myositis |
| 45543616 | Infective myositis, unspecified toe(s) | M60.078 | myositis |
| 1570735 | Other myositis, upper arm | M60.82 | myositis |
| 1570716 | Infective myositis, ankle, foot and toes | M60.07 | myositis |
| 1570740 | Other myositis, ankle and foot | M60.87 | myositis |
| 1570724 | Interstitial myositis, ankle and foot | M60.17 | myositis |
| 45538799 | Other myositis, multiple sites | M60.89 | myositis |
| 45543618 | Interstitial myositis, unspecified lower leg | M60.169 | myositis |
| 45591868 | Infective myositis, left finger(s) | M60.045 | myositis |
| 45567585 | Interstitial myositis, right upper arm | M60.121 | myositis |
| 45582178 | Interstitial myositis, unspecified shoulder | M60.119 | myositis |
| 45586995 | Infective myositis, unspecified right arm | M60.000 | myositis |
| 45577291 | Infective myositis, left lower leg | M60.062 | myositis |
| 45567589 | Other myositis, other site | M60.88 | myositis |
| 1570736 | Other myositis, forearm | M60.83 | myositis |
| 45538792 | Infective myositis, unspecified leg | M60.005 | myositis |
| 1570721 | Interstitial myositis, hand | M60.14 | myositis |
| 45553210 | Interstitial myositis, multiple sites | M60.19 | myositis |
| 45596608 | Other myositis, unspecified site | M60.80 | myositis |
| 45586997 | Interstitial myositis, unspecified ankle and foot | M60.179 | myositis |
| 45562751 | Infective myositis, unspecified hand | M60.043 | myositis |
| 45577289 | Infective myositis, right upper arm | M60.021 | myositis |
| 45577290 | Infective myositis, right hand | M60.041 | myositis |
| 45596605 | Interstitial myositis, right hand | M60.141 | myositis |
| 1570734 | Other myositis shoulder | M60.81 | myositis |
| 45533871 | Interstitial myositis, left shoulder | M60.112 | myositis |
| 45543614 | Infective myositis, unspecified shoulder | M60.019 | myositis |
| 45548451 | Other myositis, unspecified shoulder | M60.819 | myositis |
| 45596600 | Infective myositis, multiple sites | M60.09 | myositis |
| 45567587 | Other myositis, left shoulder | M60.812 | myositis |
| 45533873 | Interstitial myositis, other site | M60.18 | myositis |
| 1570717 | Interstitial myositis | M60.1 | myositis |
| 45548453 | Other myositis, right thigh | M60.851 | myositis |
| 45557912 | Other myositis, right ankle and foot | M60.871 | myositis |
| 45596604 | Interstitial myositis, unspecified forearm | M60.139 | myositis |
| 45567584 | Infective myositis, right toe(s) | M60.076 | myositis |
| 45596609 | Other myositis, unspecified upper arm | M60.829 | myositis |
| 45577288 | Infective myositis, left shoulder | M60.012 | myositis |
| 35208972 | Myositis, unspecified | M60.9 | myositis |
| 1570739 | Other myositis, lower leg | M60.86 | myositis |
| 45533872 | Interstitial myositis, left lower leg | M60.162 | myositis |
| 45596596 | Infective myositis, right shoulder | M60.011 | myositis |
| 45543615 | Infective myositis, left hand | M60.042 | myositis |
| 45596601 | Interstitial myositis, right shoulder | M60.111 | myositis |
| 1570614 | Dermatopolymyositis | M33 | myositis |
| 45596598 | Infective myositis, right finger(s) | M60.044 | myositis |
| 45596599 | Infective myositis, left thigh | M60.052 | myositis |
| 45543617 | Interstitial myositis, unspecified upper arm | M60.129 | myositis |
| 1570707 | Myositis | M60 | myositis |
| 45591869 | Infective myositis, unspecified finger(s) | M60.046 | myositis |
| 45557907 | Interstitial myositis of unspecified site | M60.10 | myositis |
| 45591871 | Interstitial myositis, left thigh | M60.152 | myositis |
| 45596597 | Infective myositis, unspecified upper arm | M60.029 | myositis |
| 45538794 | Infective myositis, left ankle | M60.071 | myositis |
| 1570737 | Other myositis, hand | M60.84 | myositis |
| 45586996 | Interstitial myositis, left upper arm | M60.122 | myositis |
| 1570710 | Infective myositis, shoulder | M60.01 | myositis |
| 45548452 | Other myositis, right forearm | M60.831 | myositis |
| 45548445 | Infective myositis, unspecified left arm | M60.001 | myositis |
| 45572472 | Infective myositis, left toe(s) | M60.077 | myositis |
| 45572475 | Other myositis, right hand | M60.841 | myositis |
| 45543621 | Other myositis, unspecified hand | M60.849 | myositis |
| 45582179 | Interstitial myositis, right thigh | M60.151 | myositis |
| 1570715 | Infective myositis, lower leg | M60.06 | myositis |
| 45572473 | Interstitial myositis, left ankle and foot | M60.172 | myositis |
| 1570733 | Other myositis | M60.8 | myositis |
| 45562752 | Infective myositis, unspecified thigh | M60.059 | myositis |
| 45577292 | Infective myositis, unspecified lower leg | M60.069 | myositis |
| 45557910 | Other myositis, right shoulder | M60.811 | myositis |
| 45548446 | Infective myositis, left foot | M60.074 | myositis |
| 45553209 | Interstitial myositis, unspecified hand | M60.149 | myositis |
| 45538798 | Other myositis, left upper arm | M60.822 | myositis |
| 1570711 | Infective myositis, upper arm | M60.02 | myositis |
| 45596602 | Interstitial myositis, right forearm | M60.131 | myositis |
| 1570723 | Interstitial myositis, lower leg | M60.16 | myositis |
| 1570719 | Interstitial myositis, upper arm | M60.12 | myositis |
| 45567583 | Infective myositis, left upper arm | M60.022 | myositis |
| 45553212 | Other myositis, left lower leg | M60.862 | myositis |
| 45572476 | Other myositis, left ankle and foot | M60.872 | myositis |
| 45533870 | Infective myositis, unspecified ankle | M60.072 | myositis |
| 1570713 | Infective myositis, hand and fingers | M60.04 | myositis |
| 45606249 | Infective myositis, right forearm | M60.031 | myositis |
| 45557911 | Other myositis, right upper arm | M60.821 | myositis |
| 1570714 | Infective myositis, thigh | M60.05 | myositis |
| 45586998 | Other myositis, left forearm | M60.832 | myositis |
| 45538793 | Infective myositis, right thigh | M60.051 | myositis |
| 45582184 | Other myositis, unspecified thigh | M60.859 | myositis |
| 45591873 | Other myositis, left thigh | M60.852 | myositis |
| 45538795 | Infective myositis, right foot | M60.073 | myositis |
| 35211275 | Cough | R05 | respiratory_signs_and_sx |
| 766424 | Acute cough | R05.1 | respiratory_signs_and_sx |
| 766427 | Cough syncope | R05.4 | respiratory_signs_and_sx |
| 766428 | Other specified cough | R05.8 | respiratory_signs_and_sx |
| 766429 | Cough, unspecified | R05.9 | respiratory_signs_and_sx |
| 766425 | Subacute cough | R05.2 | respiratory_signs_and_sx |
| 766426 | Chronic cough | R05.3 | respiratory_signs_and_sx |
| 45534422 | Shortness of breath | R06.02 | respiratory_signs_and_sx |
| 35211286 | Pleurisy | R09.1 | respiratory_signs_and_sx |
| 35211272 | Hemoptysis | R04.2 | respiratory_signs_and_sx |
| 35211278 | Periodic breathing | R06.3 | respiratory_signs_and_sx |
| 45573007 | Hypoxemia | R09.02 | respiratory_signs_and_sx |
| 35211273 | Hemorrhage from other sites in respiratory passages | R04.8 | respiratory_signs_and_sx |
| 1572196 | Asphyxia and hypoxemia | R09.0 | respiratory_signs_and_sx |
| 35211287 | Respiratory arrest | R09.2 | respiratory_signs_and_sx |
| 35211276 | Stridor | R06.1 | respiratory_signs_and_sx |
| 45558453 | Other specified symptoms and signs involving the circulatory and respiratory systems | R09.89 | respiratory_signs_and_sx |
| 45563287 | Hemorrhage from other sites in respiratory passages | R04.89 | respiratory_signs_and_sx |
| 35211274 | Hemorrhage from respiratory passages, unspecified | R04.9 | respiratory_signs_and_sx |
| 35211280 | Mouth breathing | R06.5 | respiratory_signs_and_sx |
| 35211282 | Sneezing | R06.7 | respiratory_signs_and_sx |
| 1572192 | Other abnormalities of breathing | R06.8 | respiratory_signs_and_sx |
| 45587496 | Dyspnea, unspecified | R06.00 | respiratory_signs_and_sx |
| 35211279 | Hyperventilation | R06.4 | respiratory_signs_and_sx |
| 35211277 | Wheezing | R06.2 | respiratory_signs_and_sx |
| 1572190 | Abnormalities of breathing | R06 | respiratory_signs_and_sx |
| 35211271 | Hemorrhage from throat | R04.1 | respiratory_signs_and_sx |
| 45606793 | Apnea, not elsewhere classified | R06.81 | respiratory_signs_and_sx |
| 1572189 | Hemorrhage from respiratory passages | R04 | respiratory_signs_and_sx |
| 45597165 | Orthopnea | R06.01 | respiratory_signs_and_sx |
| 35211288 | Abnormal sputum | R09.3 | respiratory_signs_and_sx |
| 45553714 | Snoring | R06.83 | respiratory_signs_and_sx |
| 45568110 | Asphyxia | R09.01 | respiratory_signs_and_sx |
| 35211270 | Epistaxis | R04.0 | respiratory_signs_and_sx |
| 1326788 | Acute respiratory distress | R06.03 | respiratory_signs_and_sx |
| 1572191 | Dyspnea | R06.0 | respiratory_signs_and_sx |
| 1572195 | Other symptoms and signs involving the circulatory and respiratory system | R09 | respiratory_signs_and_sx |
| 45539314 | Tachypnea, not elsewhere classified | R06.82 | respiratory_signs_and_sx |
| 1572197 | Other specified symptoms and signs involving the circulatory and respiratory systems | R09.8 | respiratory_signs_and_sx |
| 45563288 | Unspecified abnormalities of breathing | R06.9 | respiratory_signs_and_sx |
| 35211281 | Hiccough | R06.6 | respiratory_signs_and_sx |
| 45602003 | Nasal congestion | R09.81 | respiratory_signs_and_sx |
| 45577778 | Other abnormalities of breathing | R06.89 | respiratory_signs_and_sx |
| 45548944 | Other forms of dyspnea | R06.09 | respiratory_signs_and_sx |
| 45606794 | Postnasal drip | R09.82 | respiratory_signs_and_sx |
| 35211311 | Localized swelling, mass and lump, neck | R22.1 | skin |
| 1572214 | Localized swelling, mass and lump, upper limb | R22.3 | skin |
| 35211310 | Localized swelling, mass and lump, head | R22.0 | skin |
| 45553722 | Localized swelling, mass and lump, left upper limb | R22.32 | skin |
| 35208688 | Other atrophic disorders of skin | L90.8 | skin |
| 45568121 | Localized swelling, mass and lump, upper limb, bilateral | R22.33 | skin |
| 35208698 | Granulomatous disorder of the skin and subcutaneous tissue, unspecified | L92.9 | skin |
| 35211307 | Hyperesthesia | R20.3 | skin |
| 35208689 | Atrophic disorder of skin, unspecified | L90.9 | skin |
| 35211318 | Changes in skin texture | R23.4 | skin |
| 45606799 | Unspecified disturbances of skin sensation | R20.9 | skin |
| 45539324 | Localized swelling, mass and lump, right upper limb | R22.31 | skin |
| 1572215 | Localized swelling, mass and lump, lower limb | R22.4 | skin |
| 45544135 | Localized swelling, mass and lump, unspecified upper limb | R22.30 | skin |
| 35211306 | Paresthesia of skin | R20.2 | skin |
| 45548957 | Localized swelling, mass and lump, left lower limb | R22.42 | skin |
| 35208656 | Postinflammatory hyperpigmentation | L81.0 | skin |
| 35211309 | Rash and other nonspecific skin eruption | R21 | skin |
| 35211319 | Other skin changes | R23.8 | skin |
| 35208642 | Follicular disorder, unspecified | L73.9 | skin |
| 45573019 | Localized swelling, mass and lump, unspecified lower limb | R22.40 | skin |
| 35208672 | Other specified epidermal thickening | L85.8 | skin |
| 45548958 | Localized swelling, mass and lump, lower limb, bilateral | R22.43 | skin |
| 45606800 | Unspecified skin changes | R23.9 | skin |
| 35208709 | Other specified localized connective tissue disorders | L94.8 | skin |
| 35211308 | Other disturbances of skin sensation | R20.8 | skin |
| 45592413 | Localized swelling, mass and lump, right lower limb | R22.41 | skin |
| 1572213 | Localized swelling, mass and lump of skin and subcutaneous tissue | R22 | skin |
| 35211312 | Localized swelling, mass and lump, trunk | R22.2 | skin |
| 35208673 | Epidermal thickening, unspecified | L85.9 | skin |
| 35211316 | Flushing | R23.2 | skin |
| 35211317 | Spontaneous ecchymoses | R23.3 | skin |
| 35208720 | Other infiltrative disorders of the skin and subcutaneous tissue | L98.6 | skin |
| 1572212 | Disturbances of skin sensation | R20 | skin |
| 1572216 | Other skin changes | R23 | skin |
| 35211313 | Localized swelling, mass and lump, unspecified | R22.9 | skin |
| 35208723 | Other disorders of skin and subcutaneous tissue in diseases classified elsewhere | L99 | skin |
| 35208722 | Disorder of the skin and subcutaneous tissue, unspecified | L98.9 | skin |
| 45536984 | Chilblains, subsequent encounter | T69.1XXD | skin |
| 45599694 | Chilblains, sequela | T69.1XXS | skin |
| 45551429 | Chilblains, initial encounter | T69.1XXA | skin |
| 19160 | Chilblains | T69.1 | skin |
| 35208481 | Unspecified contact dermatitis, unspecified cause | L25.9 | skin |
| 35208496 | Pruritus, unspecified | L29.9 | skin |
| 35208495 | Other pruritus | L29.8 | skin |
| 35208510 | Other psoriasis | L40.8 | skin |
| 35208516 | Large plaque parapsoriasis | L41.4 | skin |
| 35208509 | Guttate psoriasis | L40.4 | skin |
| 35208515 | Small plaque parapsoriasis | L41.3 | skin |
| 45586768 | Psoriatic spondylitis | L40.53 | skin |
| 35208507 | Acrodermatitis continua | L40.2 | skin |
| 1569777 | Psoriasis | L40 | skin |
| 35208517 | Retiform parapsoriasis | L41.5 | skin |
| 1569779 | Parapsoriasis | L41 | skin |
| 45548197 | Psoriatic arthritis mutilans | L40.52 | skin |
| 45596373 | Other psoriatic arthropathy | L40.59 | skin |
| 35208508 | Pustulosis palmaris et plantaris | L40.3 | skin |
| 45548196 | Arthropathic psoriasis, unspecified | L40.50 | skin |
| 35208505 | Psoriasis vulgaris | L40.0 | skin |
| 45601222 | Distal interphalangeal psoriatic arthropathy | L40.51 | skin |
| 45533640 | Psoriatic juvenile arthropathy | L40.54 | skin |
| 35208511 | Psoriasis, unspecified | L40.9 | skin |
| 35208506 | Generalized pustular psoriasis | L40.1 | skin |
| 35208519 | Parapsoriasis, unspecified | L41.9 | skin |
| 1569778 | Arthropathic psoriasis | L40.5 | skin |
| 35208518 | Other parapsoriasis | L41.8 | skin |
| 35211315 | Pallor | R23.1 | skin |
| 35211305 | Hypoesthesia of skin | R20.1 | skin |
| 35211304 | Anesthesia of skin | R20.0 | skin |
| 45586344 | Vitiligo of right upper eyelid and periocular area | H02.731 | skin |
| 45552568 | Vitiligo of left lower eyelid and periocular area | H02.735 | skin |
| 45566930 | Vitiligo of right lower eyelid and periocular area | H02.732 | skin |
| 45538173 | Vitiligo of right eye, unspecified eyelid and periocular area | H02.733 | skin |
| 1568479 | Vitiligo of eyelid and periocular area | H02.73 | skin |
| 45605586 | Vitiligo of left eye, unspecified eyelid and periocular area | H02.736 | skin |
| 45557286 | Vitiligo of left upper eyelid and periocular area | H02.734 | skin |
| 45542944 | Vitiligo of unspecified eye, unspecified eyelid and periocular area | H02.739 | skin |
| 35208655 | Vitiligo | L80 | skin |
| 45543237 | Acute embolism and thrombosis of axillary vein, bilateral | I82.A13 | thrombophlebitis_and_thromboembolism |
| 45533514 | Chronic embolism and thrombosis of unspecified veins of right upper extremity | I82.701 | thrombophlebitis_and_thromboembolism |
| 45552851 | Acute embolism and thrombosis of left popliteal vein | I82.432 | thrombophlebitis_and_thromboembolism |
| 45567226 | Atheroembolism of unspecified lower extremity | I75.029 | thrombophlebitis_and_thromboembolism |
| 45557595 | Phlebitis and thrombophlebitis of popliteal vein, bilateral | I80.223 | thrombophlebitis_and_thromboembolism |
| 45534180 | Deep phlebothrombosis in pregnancy, unspecified trimester | O22.30 | thrombophlebitis_and_thromboembolism |
| 45543223 | Phlebitis and thrombophlebitis of other deep vessels of right lower extremity | I80.291 | thrombophlebitis_and_thromboembolism |
| 45562409 | Acute embolism and thrombosis of unspecified veins of unspecified upper extremity | I82.609 | thrombophlebitis_and_thromboembolism |
| 45572138 | Acute embolism and thrombosis of unspecified deep veins of left proximal lower extremity | I82.4Y2 | thrombophlebitis_and_thromboembolism |
| 45596248 | Other arterial embolism and thrombosis of abdominal aorta | I74.09 | thrombophlebitis_and_thromboembolism |
| 35207862 | Embolism and thrombosis of arteries of the upper extremities | I74.2 | thrombophlebitis_and_thromboembolism |
| 45601093 | Acute embolism and thrombosis of superficial veins of upper extremity, bilateral | I82.613 | thrombophlebitis_and_thromboembolism |
| 45533513 | Chronic embolism and thrombosis of unspecified deep veins of unspecified distal lower extremity | I82.5Z9 | thrombophlebitis_and_thromboembolism |
| 35207886 | Phlebitis and thrombophlebitis of unspecified site | I80.9 | thrombophlebitis_and_thromboembolism |
| 45596255 | Chronic embolism and thrombosis of unspecified popliteal vein | I82.539 | thrombophlebitis_and_thromboembolism |
| 45543217 | Atheroembolism of right lower extremity | I75.021 | thrombophlebitis_and_thromboembolism |
| 45591522 | Chronic embolism and thrombosis of left axillary vein | I82.A22 | thrombophlebitis_and_thromboembolism |
| 45605849 | Acute embolism and thrombosis of iliac vein, bilateral | I82.423 | thrombophlebitis_and_thromboembolism |
| 45533507 | Phlebitis and thrombophlebitis of left popliteal vein | I80.222 | thrombophlebitis_and_thromboembolism |
| 45581821 | Acute embolism and thrombosis of tibial vein, bilateral | I82.443 | thrombophlebitis_and_thromboembolism |
| 45576928 | Chronic embolism and thrombosis of other specified veins | I82.891 | thrombophlebitis_and_thromboembolism |
| 45572083 | Septic pulmonary embolism with acute cor pulmonale | I26.01 | thrombophlebitis_and_thromboembolism |
| 45538452 | Chronic embolism and thrombosis of superior vena cava | I82.211 | thrombophlebitis_and_thromboembolism |
| 45586635 | Acute embolism and thrombosis of left femoral vein | I82.412 | thrombophlebitis_and_thromboembolism |
| 35207863 | Embolism and thrombosis of arteries of the lower extremities | I74.3 | thrombophlebitis_and_thromboembolism |
| 1553776 | Chronic embolism and thrombosis of right peroneal vein | I82.551 | thrombophlebitis_and_thromboembolism |
| 45548082 | Acute embolism and thrombosis of deep veins of unspecified upper extremity | I82.629 | thrombophlebitis_and_thromboembolism |
| 45538454 | Acute embolism and thrombosis of left iliac vein | I82.422 | thrombophlebitis_and_thromboembolism |
| 45538456 | Acute embolism and thrombosis of deep veins of left upper extremity | I82.622 | thrombophlebitis_and_thromboembolism |
| 45562406 | Acute embolism and thrombosis of unspecified tibial vein | I82.449 | thrombophlebitis_and_thromboembolism |
| 45543229 | Acute embolism and thrombosis of unspecified deep veins of right lower extremity | I82.401 | thrombophlebitis_and_thromboembolism |
| 45605853 | Chronic embolism and thrombosis of unspecified femoral vein | I82.519 | thrombophlebitis_and_thromboembolism |
| 45576917 | Saddle embolus of abdominal aorta | I74.01 | thrombophlebitis_and_thromboembolism |
| 45591521 | Chronic embolism and thrombosis of unspecified deep veins of left distal lower extremity | I82.5Z2 | thrombophlebitis_and_thromboembolism |
| 45596257 | Chronic embolism and thrombosis of superficial veins of unspecified upper extremity | I82.719 | thrombophlebitis_and_thromboembolism |
| 45567234 | Acute embolism and thrombosis of unspecified deep veins of unspecified lower extremity | I82.409 | thrombophlebitis_and_thromboembolism |
| 45591518 | Phlebitis and thrombophlebitis of superficial vessels of right lower extremity | I80.01 | thrombophlebitis_and_thromboembolism |
| 45576925 | Chronic embolism and thrombosis of unspecified deep veins of unspecified proximal lower extremity | I82.5Y9 | thrombophlebitis_and_thromboembolism |
| 1553756 | Phlebitis and thrombophlebitis of right peroneal vein | I80.241 | thrombophlebitis_and_thromboembolism |
| 45581823 | Chronic embolism and thrombosis of unspecified deep veins of left proximal lower extremity | I82.5Y2 | thrombophlebitis_and_thromboembolism |
| 45586576 | Other pulmonary embolism with acute cor pulmonale | I26.09 | thrombophlebitis_and_thromboembolism |
| 45543234 | Chronic embolism and thrombosis of superficial veins of left upper extremity | I82.712 | thrombophlebitis_and_thromboembolism |
| 45572143 | Chronic embolism and thrombosis of other specified deep vein of unspecified lower extremity | I82.599 | thrombophlebitis_and_thromboembolism |
| 45557593 | Phlebitis and thrombophlebitis of unspecified femoral vein | I80.10 | thrombophlebitis_and_thromboembolism |
| 45533510 | Acute embolism and thrombosis of other specified deep vein of right lower extremity | I82.491 | thrombophlebitis_and_thromboembolism |
| 45587287 | Deep phlebothrombosis in pregnancy, first trimester | O22.31 | thrombophlebitis_and_thromboembolism |
| 1553769 | Acute embolism and thrombosis of unspecified peroneal vein | I82.459 | thrombophlebitis_and_thromboembolism |
| 45572144 | Acute embolism and thrombosis of superficial veins of unspecified upper extremity | I82.619 | thrombophlebitis_and_thromboembolism |
| 45605846 | Phlebitis and thrombophlebitis of right tibial vein | I80.231 | thrombophlebitis_and_thromboembolism |
| 45605789 | Septic pulmonary embolism without acute cor pulmonale | I26.90 | thrombophlebitis_and_thromboembolism |
| 45605856 | Chronic embolism and thrombosis of deep veins of right upper extremity | I82.721 | thrombophlebitis_and_thromboembolism |
| 45543222 | Phlebitis and thrombophlebitis of unspecified tibial vein | I80.239 | thrombophlebitis_and_thromboembolism |
| 45581820 | Acute embolism and thrombosis of right femoral vein | I82.411 | thrombophlebitis_and_thromboembolism |
| 45606541 | Superficial thrombophlebitis in pregnancy, second trimester | O22.22 | thrombophlebitis_and_thromboembolism |
| 45557594 | Phlebitis and thrombophlebitis of unspecified iliac vein | I80.219 | thrombophlebitis_and_thromboembolism |
| 45538450 | Phlebitis and thrombophlebitis of right popliteal vein | I80.221 | thrombophlebitis_and_thromboembolism |
| 45538445 | Embolism and thrombosis of unspecified parts of aorta | I74.10 | thrombophlebitis_and_thromboembolism |
| 1553749 | Single subsegmental pulmonary embolism without acute cor pulmonale | I26.93 | thrombophlebitis_and_thromboembolism |
| 45533441 | Chronic pulmonary embolism | I27.82 | thrombophlebitis_and_thromboembolism |
| 45567240 | Chronic embolism and thrombosis of unspecified veins of upper extremity, bilateral | I82.703 | thrombophlebitis_and_thromboembolism |
| 35210369 | Superficial thrombophlebitis in the puerperium | O87.0 | thrombophlebitis_and_thromboembolism |
| 45576924 | Chronic embolism and thrombosis of unspecified tibial vein | I82.549 | thrombophlebitis_and_thromboembolism |
| 45572135 | Phlebitis and thrombophlebitis of unspecified deep vessels of right lower extremity | I80.201 | thrombophlebitis_and_thromboembolism |
| 45605857 | Embolism and thrombosis of superficial veins of lower extremities, bilateral | I82.813 | thrombophlebitis_and_thromboembolism |
| 45591523 | Acute embolism and thrombosis of unspecified subclavian vein | I82.B19 | thrombophlebitis_and_thromboembolism |
| 45601085 | Phlebitis and thrombophlebitis of superficial vessels of lower extremities, bilateral | I80.03 | thrombophlebitis_and_thromboembolism |
| 1553763 | Phlebitis and thrombophlebitis of calf muscular vein, bilateral | I80.253 | thrombophlebitis_and_thromboembolism |
| 1553773 | Acute embolism and thrombosis of calf muscular vein, bilateral | I82.463 | thrombophlebitis_and_thromboembolism |
| 45567244 | Chronic embolism and thrombosis of subclavian vein, bilateral | I82.B23 | thrombophlebitis_and_thromboembolism |
| 35207888 | Budd-Chiari syndrome | I82.0 | thrombophlebitis_and_thromboembolism |
| 35207884 | Phlebitis and thrombophlebitis of lower extremities, unspecified | I80.3 | thrombophlebitis_and_thromboembolism |
| 45543230 | Chronic embolism and thrombosis of left femoral vein | I82.512 | thrombophlebitis_and_thromboembolism |
| 45548085 | Chronic embolism and thrombosis of unspecified axillary vein | I82.A29 | thrombophlebitis_and_thromboembolism |
| 35207889 | Thrombophlebitis migrans | I82.1 | thrombophlebitis_and_thromboembolism |
| 45533508 | Acute embolism and thrombosis of femoral vein, bilateral | I82.413 | thrombophlebitis_and_thromboembolism |
| 1569147 | Pulmonary embolism | I26 | thrombophlebitis_and_thromboembolism |
| 1553759 | Phlebitis and thrombophlebitis of unspecified peroneal vein | I80.249 | thrombophlebitis_and_thromboembolism |
| 45538449 | Phlebitis and thrombophlebitis of right femoral vein | I80.11 | thrombophlebitis_and_thromboembolism |
| 45552852 | Acute embolism and thrombosis of left tibial vein | I82.442 | thrombophlebitis_and_thromboembolism |
| 1569327 | Arterial embolism and thrombosis | I74 | thrombophlebitis_and_thromboembolism |
| 45562410 | Acute embolism and thrombosis of left subclavian vein | I82.B12 | thrombophlebitis_and_thromboembolism |
| 45576929 | Acute embolism and thrombosis of subclavian vein, bilateral | I82.B13 | thrombophlebitis_and_thromboembolism |
| 1553761 | Phlebitis and thrombophlebitis of right calf muscular vein | I80.251 | thrombophlebitis_and_thromboembolism |
| 45572084 | Other pulmonary embolism without acute cor pulmonale | I26.99 | thrombophlebitis_and_thromboembolism |
| 45576922 | Acute embolism and thrombosis of unspecified deep veins of lower extremity, bilateral | I82.403 | thrombophlebitis_and_thromboembolism |
| 45543236 | Embolism and thrombosis of superficial veins of right lower extremity | I82.811 | thrombophlebitis_and_thromboembolism |
| 45538455 | Acute embolism and thrombosis of other specified deep vein of unspecified lower extremity | I82.499 | thrombophlebitis_and_thromboembolism |
| 35207865 | Embolism and thrombosis of iliac artery | I74.5 | thrombophlebitis_and_thromboembolism |
| 1553766 | Acute embolism and thrombosis of right peroneal vein | I82.451 | thrombophlebitis_and_thromboembolism |
| 45543224 | Phlebitis and thrombophlebitis of other deep vessels of left lower extremity | I80.292 | thrombophlebitis_and_thromboembolism |
| 45601094 | Acute embolism and thrombosis of deep veins of right upper extremity | I82.621 | thrombophlebitis_and_thromboembolism |
| 45601095 | Chronic embolism and thrombosis of unspecified subclavian vein | I82.B29 | thrombophlebitis_and_thromboembolism |
| 45567241 | Embolism and thrombosis of superficial veins of left lower extremity | I82.812 | thrombophlebitis_and_thromboembolism |
| 45581825 | Acute embolism and thrombosis of unspecified veins of left upper extremity | I82.602 | thrombophlebitis_and_thromboembolism |
| 1553781 | Chronic embolism and thrombosis of right calf muscular vein | I82.561 | thrombophlebitis_and_thromboembolism |
| 45567238 | Chronic embolism and thrombosis of left iliac vein | I82.522 | thrombophlebitis_and_thromboembolism |
| 45533506 | Phlebitis and thrombophlebitis of superficial vessels of left lower extremity | I80.02 | thrombophlebitis_and_thromboembolism |
| 1553762 | Phlebitis and thrombophlebitis of left calf muscular vein | I80.252 | thrombophlebitis_and_thromboembolism |
| 45543225 | Phlebitis and thrombophlebitis of other deep vessels of lower extremity, bilateral | I80.293 | thrombophlebitis_and_thromboembolism |
| 45558205 | Superficial thrombophlebitis in pregnancy, unspecified trimester | O22.20 | thrombophlebitis_and_thromboembolism |
| 1553772 | Acute embolism and thrombosis of left calf muscular vein | I82.462 | thrombophlebitis_and_thromboembolism |
| 45533518 | Embolism and thrombosis of superficial veins of unspecified lower extremity | I82.819 | thrombophlebitis_and_thromboembolism |
| 45557540 | Saddle embolus of pulmonary artery with acute cor pulmonale | I26.02 | thrombophlebitis_and_thromboembolism |
| 45543238 | Chronic embolism and thrombosis of axillary vein, bilateral | I82.A23 | thrombophlebitis_and_thromboembolism |
| 45557599 | Chronic embolism and thrombosis of deep veins of left upper extremity | I82.722 | thrombophlebitis_and_thromboembolism |
| 45562408 | Chronic embolism and thrombosis of right tibial vein | I82.541 | thrombophlebitis_and_thromboembolism |
| 1553758 | Phlebitis and thrombophlebitis of peroneal vein, bilateral | I80.243 | thrombophlebitis_and_thromboembolism |
| 45562403 | Phlebitis and thrombophlebitis of unspecified deep vessels of unspecified lower extremity | I80.209 | thrombophlebitis_and_thromboembolism |
| 45557600 | Acute embolism and thrombosis of unspecified axillary vein | I82.A19 | thrombophlebitis_and_thromboembolism |
| 35207864 | Embolism and thrombosis of arteries of extremities, unspecified | I74.4 | thrombophlebitis_and_thromboembolism |
| 45543226 | Phlebitis and thrombophlebitis of other deep vessels of unspecified lower extremity | I80.299 | thrombophlebitis_and_thromboembolism |
| 45533515 | Chronic embolism and thrombosis of unspecified veins of left upper extremity | I82.702 | thrombophlebitis_and_thromboembolism |
| 45567227 | Septic arterial embolism | I76 | thrombophlebitis_and_thromboembolism |
| 35210370 | Deep phlebothrombosis in the puerperium | O87.1 | thrombophlebitis_and_thromboembolism |
| 45539096 | Superficial thrombophlebitis in pregnancy, third trimester | O22.23 | thrombophlebitis_and_thromboembolism |
| 45552847 | Atheroembolism of kidney | I75.81 | thrombophlebitis_and_thromboembolism |
| 45562411 | Chronic embolism and thrombosis of left subclavian vein | I82.B22 | thrombophlebitis_and_thromboembolism |
| 45605852 | Chronic embolism and thrombosis of femoral vein, bilateral | I82.513 | thrombophlebitis_and_thromboembolism |
| 1553778 | Chronic embolism and thrombosis of peroneal vein, bilateral | I82.553 | thrombophlebitis_and_thromboembolism |
| 1553774 | Acute embolism and thrombosis of unspecified calf muscular vein | I82.469 | thrombophlebitis_and_thromboembolism |
| 45548081 | Acute embolism and thrombosis of unspecified veins of right upper extremity | I82.601 | thrombophlebitis_and_thromboembolism |
| 45543233 | Acute embolism and thrombosis of deep veins of upper extremity, bilateral | I82.623 | thrombophlebitis_and_thromboembolism |
| 1553768 | Acute embolism and thrombosis of peroneal vein, bilateral | I82.453 | thrombophlebitis_and_thromboembolism |
| 45605850 | Acute embolism and thrombosis of unspecified deep veins of unspecified proximal lower extremity | I82.4Y9 | thrombophlebitis_and_thromboembolism |
| 45533511 | Chronic embolism and thrombosis of other specified deep vein of right lower extremity | I82.591 | thrombophlebitis_and_thromboembolism |
| 45572141 | Chronic embolism and thrombosis of left popliteal vein | I82.532 | thrombophlebitis_and_thromboembolism |
| 45581822 | Chronic embolism and thrombosis of right femoral vein | I82.511 | thrombophlebitis_and_thromboembolism |
| 45567232 | Phlebitis and thrombophlebitis of left tibial vein | I80.232 | thrombophlebitis_and_thromboembolism |
| 45581824 | Chronic embolism and thrombosis of unspecified deep veins of distal lower extremity, bilateral | I82.5Z3 | thrombophlebitis_and_thromboembolism |
| 45591524 | Acute embolism and thrombosis of left internal jugular vein | I82.C12 | thrombophlebitis_and_thromboembolism |
| 45601089 | Acute embolism and thrombosis of unspecified deep veins of right proximal lower extremity | I82.4Y1 | thrombophlebitis_and_thromboembolism |
| 45552854 | Chronic embolism and thrombosis of right internal jugular vein | I82.C21 | thrombophlebitis_and_thromboembolism |
| 45605858 | Acute embolism and thrombosis of unspecified internal jugular vein | I82.C19 | thrombophlebitis_and_thromboembolism |
| 35207885 | Phlebitis and thrombophlebitis of other sites | I80.8 | thrombophlebitis_and_thromboembolism |
| 45605854 | Chronic embolism and thrombosis of unspecified iliac vein | I82.529 | thrombophlebitis_and_thromboembolism |
| 45596259 | Acute embolism and thrombosis of internal jugular vein, bilateral | I82.C13 | thrombophlebitis_and_thromboembolism |
| 35207797 | Intracardiac thrombosis, not elsewhere classified | I51.3 | thrombophlebitis_and_thromboembolism |
| 45577563 | Deep phlebothrombosis in pregnancy, second trimester | O22.32 | thrombophlebitis_and_thromboembolism |
| 45552853 | Chronic embolism and thrombosis of right axillary vein | I82.A21 | thrombophlebitis_and_thromboembolism |
| 45597038 | Puerperal septic thrombophlebitis | O86.81 | thrombophlebitis_and_thromboembolism |
| 45548083 | Chronic embolism and thrombosis of superficial veins of upper extremity, bilateral | I82.713 | thrombophlebitis_and_thromboembolism |
| 35207866 | Embolism and thrombosis of other arteries | I74.8 | thrombophlebitis_and_thromboembolism |
| 45543231 | Chronic embolism and thrombosis of right popliteal vein | I82.531 | thrombophlebitis_and_thromboembolism |
| 45567242 | Chronic embolism and thrombosis of unspecified vein | I82.91 | thrombophlebitis_and_thromboembolism |
| 45576921 | Phlebitis and thrombophlebitis of unspecified popliteal vein | I80.229 | thrombophlebitis_and_thromboembolism |
| 45601088 | Chronic embolism and thrombosis of inferior vena cava | I82.221 | thrombophlebitis_and_thromboembolism |
| 45576926 | Acute embolism and thrombosis of superficial veins of left upper extremity | I82.612 | thrombophlebitis_and_thromboembolism |
| 45567239 | Chronic embolism and thrombosis of unspecified deep veins of right proximal lower extremity | I82.5Y1 | thrombophlebitis_and_thromboembolism |
| 45538451 | Acute embolism and thrombosis of superior vena cava | I82.210 | thrombophlebitis_and_thromboembolism |
| 45596256 | Chronic embolism and thrombosis of unspecified veins of unspecified upper extremity | I82.709 | thrombophlebitis_and_thromboembolism |
| 45576923 | Acute embolism and thrombosis of unspecified deep veins of proximal lower extremity, bilateral | I82.4Y3 | thrombophlebitis_and_thromboembolism |
| 45605851 | Acute embolism and thrombosis of unspecified deep veins of distal lower extremity, bilateral | I82.4Z3 | thrombophlebitis_and_thromboembolism |
| 45586634 | Acute embolism and thrombosis of other thoracic veins | I82.290 | thrombophlebitis_and_thromboembolism |
| 45591525 | Chronic embolism and thrombosis of internal jugular vein, bilateral | I82.C23 | thrombophlebitis_and_thromboembolism |
| 45572140 | Chronic embolism and thrombosis of unspecified deep veins of lower extremity, bilateral | I82.503 | thrombophlebitis_and_thromboembolism |
| 1569148 | Pulmonary embolism with acute cor pulmonale | I26.0 | thrombophlebitis_and_thromboembolism |
| 45562412 | Chronic embolism and thrombosis of left internal jugular vein | I82.C22 | thrombophlebitis_and_thromboembolism |
| 45596252 | Acute embolism and thrombosis of right tibial vein | I82.441 | thrombophlebitis_and_thromboembolism |
| 45596258 | Acute embolism and thrombosis of left axillary vein | I82.A12 | thrombophlebitis_and_thromboembolism |
| 35207891 | Embolism and thrombosis of renal vein | I82.3 | thrombophlebitis_and_thromboembolism |
| 45548080 | Acute embolism and thrombosis of right iliac vein | I82.421 | thrombophlebitis_and_thromboembolism |
| 45562404 | Acute embolism and thrombosis of inferior vena cava | I82.220 | thrombophlebitis_and_thromboembolism |
| 35207867 | Embolism and thrombosis of unspecified artery | I74.9 | thrombophlebitis_and_thromboembolism |
| 45533512 | Chronic embolism and thrombosis of unspecified deep veins of right distal lower extremity | I82.5Z1 | thrombophlebitis_and_thromboembolism |
| 45576920 | Phlebitis and thrombophlebitis of unspecified deep vessels of lower extremities, bilateral | I80.203 | thrombophlebitis_and_thromboembolism |
| 1553767 | Acute embolism and thrombosis of left peroneal vein | I82.452 | thrombophlebitis_and_thromboembolism |
| 1569149 | Pulmonary embolism without acute cor pulmonale | I26.9 | thrombophlebitis_and_thromboembolism |
| 1553779 | Chronic embolism and thrombosis of unspecified peroneal vein | I82.559 | thrombophlebitis_and_thromboembolism |
| 45567224 | Atheroembolism of bilateral upper extremities | I75.013 | thrombophlebitis_and_thromboembolism |
| 45601091 | Acute embolism and thrombosis of unspecified veins of upper extremity, bilateral | I82.603 | thrombophlebitis_and_thromboembolism |
| 45581817 | Phlebitis and thrombophlebitis of right iliac vein | I80.211 | thrombophlebitis_and_thromboembolism |
| 45591520 | Chronic embolism and thrombosis of other specified deep vein of lower extremity, bilateral | I82.593 | thrombophlebitis_and_thromboembolism |
| 35207887 | Portal vein thrombosis | I81 | thrombophlebitis_and_thromboembolism |
| 45533519 | Chronic embolism and thrombosis of right subclavian vein | I82.B21 | thrombophlebitis_and_thromboembolism |
| 45562405 | Acute embolism and thrombosis of right popliteal vein | I82.431 | thrombophlebitis_and_thromboembolism |
| 45596253 | Acute embolism and thrombosis of other specified deep vein of left lower extremity | I82.492 | thrombophlebitis_and_thromboembolism |
| 35207861 | Embolism and thrombosis of abdominal aorta | I74.0 | thrombophlebitis_and_thromboembolism |
| 45576918 | Atheroembolism of unspecified upper extremity | I75.019 | thrombophlebitis_and_thromboembolism |
| 45538457 | Chronic embolism and thrombosis of unspecified internal jugular vein | I82.C29 | thrombophlebitis_and_thromboembolism |
| 1569328 | Embolism and thrombosis of other and unspecified parts of aorta | I74.1 | thrombophlebitis_and_thromboembolism |
| 45567236 | Chronic embolism and thrombosis of unspecified deep veins of right lower extremity | I82.501 | thrombophlebitis_and_thromboembolism |
| 45533516 | Chronic embolism and thrombosis of superficial veins of right upper extremity | I82.711 | thrombophlebitis_and_thromboembolism |
| 35207696 | Thrombosis of atrium, auricular appendage, and ventricle as current complications following acute myocardial infarction | I23.6 | thrombophlebitis_and_thromboembolism |
| 45596254 | Chronic embolism and thrombosis of right iliac vein | I82.521 | thrombophlebitis_and_thromboembolism |
| 45601083 | Atheroembolism of right upper extremity | I75.011 | thrombophlebitis_and_thromboembolism |
| 45592192 | Deep phlebothrombosis in pregnancy, third trimester | O22.33 | thrombophlebitis_and_thromboembolism |
| 45586636 | Acute embolism and thrombosis of unspecified deep veins of unspecified distal lower extremity | I82.4Z9 | thrombophlebitis_and_thromboembolism |
| 45591515 | Atheroembolism of other site | I75.89 | thrombophlebitis_and_thromboembolism |
| 45596251 | Acute embolism and thrombosis of popliteal vein, bilateral | I82.433 | thrombophlebitis_and_thromboembolism |
| 1553784 | Chronic embolism and thrombosis of unspecified calf muscular vein | I82.569 | thrombophlebitis_and_thromboembolism |
| 45533517 | Chronic embolism and thrombosis of deep veins of unspecified upper extremity | I82.729 | thrombophlebitis_and_thromboembolism |
| 1553782 | Chronic embolism and thrombosis of left calf muscular vein | I82.562 | thrombophlebitis_and_thromboembolism |
| 45543232 | Chronic embolism and thrombosis of other specified deep vein of left lower extremity | I82.592 | thrombophlebitis_and_thromboembolism |
| 1553764 | Phlebitis and thrombophlebitis of unspecified calf muscular vein | I80.259 | thrombophlebitis_and_thromboembolism |
| 45552786 | Saddle embolus of pulmonary artery without acute cor pulmonale | I26.92 | thrombophlebitis_and_thromboembolism |
| 45567243 | Acute embolism and thrombosis of right subclavian vein | I82.B11 | thrombophlebitis_and_thromboembolism |
| 45533501 | Atheroembolism of left upper extremity | I75.012 | thrombophlebitis_and_thromboembolism |
| 45543235 | Chronic embolism and thrombosis of deep veins of upper extremity, bilateral | I82.723 | thrombophlebitis_and_thromboembolism |
| 45601086 | Phlebitis and thrombophlebitis of femoral vein, bilateral | I80.13 | thrombophlebitis_and_thromboembolism |
| 45557598 | Chronic embolism and thrombosis of tibial vein, bilateral | I82.543 | thrombophlebitis_and_thromboembolism |
| 45572142 | Chronic embolism and thrombosis of left tibial vein | I82.542 | thrombophlebitis_and_thromboembolism |
| 45533520 | Acute embolism and thrombosis of right internal jugular vein | I82.C11 | thrombophlebitis_and_thromboembolism |
| 1553783 | Chronic embolism and thrombosis of calf muscular vein, bilateral | I82.563 | thrombophlebitis_and_thromboembolism |
| 45605848 | Acute embolism and thrombosis of unspecified femoral vein | I82.419 | thrombophlebitis_and_thromboembolism |
| 45601092 | Acute embolism and thrombosis of superficial veins of right upper extremity | I82.611 | thrombophlebitis_and_thromboembolism |
| 45591519 | Phlebitis and thrombophlebitis of left femoral vein | I80.12 | thrombophlebitis_and_thromboembolism |
| 45557590 | Embolism and thrombosis of other parts of aorta | I74.19 | thrombophlebitis_and_thromboembolism |
| 45572146 | Acute embolism and thrombosis of right axillary vein | I82.A11 | thrombophlebitis_and_thromboembolism |
| 45581819 | Chronic embolism and thrombosis of other thoracic veins | I82.291 | thrombophlebitis_and_thromboembolism |
| 45538446 | Embolism and thrombosis of thoracic aorta | I74.11 | thrombophlebitis_and_thromboembolism |
| 45548084 | Acute embolism and thrombosis of unspecified vein | I82.90 | thrombophlebitis_and_thromboembolism |
| 45562407 | Chronic embolism and thrombosis of iliac vein, bilateral | I82.523 | thrombophlebitis_and_thromboembolism |
| 45586632 | Phlebitis and thrombophlebitis of superficial vessels of unspecified lower extremity | I80.00 | thrombophlebitis_and_thromboembolism |
| 45601090 | Chronic embolism and thrombosis of popliteal vein, bilateral | I82.533 | thrombophlebitis_and_thromboembolism |
| 45605855 | Chronic embolism and thrombosis of unspecified deep veins of proximal lower extremity, bilateral | I82.5Y3 | thrombophlebitis_and_thromboembolism |
| 1553771 | Acute embolism and thrombosis of right calf muscular vein | I82.461 | thrombophlebitis_and_thromboembolism |
| 1553757 | Phlebitis and thrombophlebitis of left peroneal vein | I80.242 | thrombophlebitis_and_thromboembolism |
| 1553777 | Chronic embolism and thrombosis of left peroneal vein | I82.552 | thrombophlebitis_and_thromboembolism |
| 45586637 | Chronic embolism and thrombosis of unspecified deep veins of left lower extremity | I82.502 | thrombophlebitis_and_thromboembolism |
| 45596250 | Acute embolism and thrombosis of unspecified iliac vein | I82.429 | thrombophlebitis_and_thromboembolism |
| 45534178 | Superficial thrombophlebitis in pregnancy, first trimester | O22.21 | thrombophlebitis_and_thromboembolism |
| 45557597 | Acute embolism and thrombosis of unspecified deep veins of right distal lower extremity | I82.4Z1 | thrombophlebitis_and_thromboembolism |
| 45548079 | Phlebitis and thrombophlebitis of unspecified deep vessels of left lower extremity | I80.202 | thrombophlebitis_and_thromboembolism |
| 45586627 | Atheroembolism of left lower extremity | I75.022 | thrombophlebitis_and_thromboembolism |
| 45538453 | Acute embolism and thrombosis of unspecified deep veins of left lower extremity | I82.402 | thrombophlebitis_and_thromboembolism |
| 1553750 | Multiple subsegmental pulmonary emboli without acute cor pulmonale | I26.94 | thrombophlebitis_and_thromboembolism |
| 45576927 | Acute embolism and thrombosis of other specified veins | I82.890 | thrombophlebitis_and_thromboembolism |
| 45567237 | Chronic embolism and thrombosis of unspecified deep veins of unspecified lower extremity | I82.509 | thrombophlebitis_and_thromboembolism |
| 45533509 | Acute embolism and thrombosis of unspecified popliteal vein | I82.439 | thrombophlebitis_and_thromboembolism |
| 45572139 | Acute embolism and thrombosis of unspecified deep veins of left distal lower extremity | I82.4Z2 | thrombophlebitis_and_thromboembolism |
| 45572136 | Phlebitis and thrombophlebitis of tibial vein, bilateral | I80.233 | thrombophlebitis_and_thromboembolism |
| 45567225 | Atheroembolism of bilateral lower extremities | I75.023 | thrombophlebitis_and_thromboembolism |
| 45543221 | Phlebitis and thrombophlebitis of left iliac vein | I80.212 | thrombophlebitis_and_thromboembolism |
| 45567231 | Phlebitis and thrombophlebitis of iliac vein, bilateral | I80.213 | thrombophlebitis_and_thromboembolism |
| 45567235 | Acute embolism and thrombosis of other specified deep vein of lower extremity, bilateral | I82.493 | thrombophlebitis_and_thromboembolism |
| 1572254 | Fever of other and unknown origin | R50 | fever_and_chills |
| 35211385 | Drug induced fever | R50.2 | fever_and_chills |
| 35211386 | Other specified fever | R50.8 | fever_and_chills |
| 45606818 | Fever presenting with conditions classified elsewhere | R50.81 | fever_and_chills |
| 45597189 | Postprocedural fever | R50.82 | fever_and_chills |
| 45592424 | Postvaccination fever | R50.83 | fever_and_chills |
| 45597190 | Febrile nonhemolytic transfusion reaction | R50.84 | fever_and_chills |
| 35211387 | Fever, unspecified | R50.9 | fever_and_chills |
| 45577807 | Chills (without fever) | R68.83 | fever_and_chills |
| 35205632 | Relapsing fever, unspecified | A68.9 | fever_and_chills |
| 35211414 | Hypothermia, not associated with low environmental temperature | R68.0 | fever_and_chills |
