## Supplementary material for "Vaccine Effectiveness Against Long COVID in Children: A Report from the RECOVER EHR Cohort": RECOVER Consortium

**Consortium Members****hip**

### **Adult Cohort**

**Brigham and Women's Hospital (BWH)**

Brigham and Women's Hospital (BWH)

Massachusetts General Hospital (MGH)

Beth Israel Deaconess Medical Center (BIDMC)

Boston University/Boston Medical Center (BMC)

Cambridge Health Alliance (CHA)

Tufts Medical Center (TMC)

South Shore Hospital

**Case Western Reserve University**

Case Western Reserve University/University Hospital

The MetroHealth System (MHS)

**Columbia University (C4R) - C4R Sites to be removed.**

CUIMC/NOMAS Cohort

**Emory**

Emory Healthcare (Hope Clinic)

Grady Health System

Morehouse School of Medicine

Atlanta VA Health Care System

Kaiser Permanente of Georgia (KPGA)

**Howard University**

Howard University

University of Maryland Mercy Medical Center (UMD-MC)

**Icahn School of Medicine at Mount Sinai**

Icahn School of Medicine at Mount Sinai

**Stanford University**

Stanford University

Stanford Tri Valley

**Institute for Systems Biology**

Swedish Health Services/ISB/Swedish Medical Center

University of Washington

Providence Regional Medical Center Everett

Providence Sacred Heart Medical Center

Cedars-Sinai Medical Center - Los Angeles, CA

**University of Alabama at Birmingham (UAB)**

University of Alabama at Birmingham (UAB)

University of South Alabama (USA)

University Medical Center New Orleans

**University of Arizona**

University of Arizona

Banner University Medical Center (BUMC) Phoenix

Banner University Medical Center (BUMC) Tucson

**University of California San Francisco**

UCSF San Francisco General Hospital (SFGH)

Parnassus Medical Center (UCSF)

San Mateo County Health Department

**University of Illinois at Chicago**

University of Illinois at Chicago

Mile Square Health Centers

BrightStar Community Outreach

Illinois Unidos

Peoria City/County Health Department

**University of Texas Health Science Center at San Antonio**

University of Texas Health Science Center at San Antonio

UT Education and Research Center at Laredo

**University of Utah**

University of Utah

Intermountain Healthcare

University of Colorado Anschutz Medical Campus

Denver Health and Hospital Authority

University of New Mexico Health Sciences Center

**West Virginia University**

West Virginia IDeA CTR

Sanford Health

University of Hawaii/Mountain West CTR

University of Kansas Medical Center

University of Kentucky

MaineHealth/Northern New England IDeA CTR (NNE-CTR

LSU Health Sciences Center New Orleans (Louisiana

Tulane School of Medicine

LSU Pennington Biomedical Research Center (Louisi

University of Mississippi Medical Center

University of Nebraska Medical Center

University of Oklahoma Health Sciences Center

Hispanic Alliance for Clinical and Translational Research

### **Adult Pregnancy Cohort**

**University of California San Francisco**

University of California San Francisco (Pregnancy)

**University of Utah**

University of Utah (Pregnancy)

University of Alabama at Birmingham

University of Texas HSC at Houston - Memorial Hermann Texas Medical Center

University of Texas Medical Branch at Galveston

UH MacDonald's Women's Hospital

Yale University

University of Pittsburgh

University of Pennsylvania

University of North Carolina - Chapel Hill

Saint Peter's University Hospital

Ohio State University

Northwestern University

NorthShore University HealthSystem

New York-Presbyterian/Queens

Miami Valley Hospital

Case Western - MetroHealth Medical

Medical College of Wisconsin

Good Samaritan

Duke University Medical Center

ChristianaCare

University of Texas HSC at Houston - Memorial City

Brown University (Women and Infants Hospital)

University of Texas HSC at Houston - LBJ Hospital

Columbia University

### **Pediatric Cohort**

**Arkansas Children's Research Institute**

University of New Mexico Health Sciences Center

West Virginia University

Pennington Biomedical Research Center

Arkansas Children's Research Institute

Nemours Children's Health System

Kapiolani Medical Center for Women and Children

University of Louisville Research Foundation, North

University of Nebraska Medical Center, Children's

Dartmouth Hitchcock Medical Center

University of Oklahoma Health Sciences Center

Northeastern University, Puerto Rico Testsite for

Medical University of South Carolina

Avera Research Institute

University of Vermont Medical Center

**Children's Hospital Los Angeles**

Children's Hospital Los Angeles

**Healthcore, NERI, MUSIC**

Children's Healthcare of Atlanta

Medical University of South Carolina

Primary Children's Hospital/University of Utah

Baylor/Texas Children's Hospital

Boston Children's Hospital

Children's Hospital of Colorado

Children's Hospital Los Angeles

Children's Hospital of Michigan

Children's Hospital of New Orleans

Children's Hospital of Philadelphia

Children's Mercy Hospital

Children's National Hospital

Cincinnati Children's Hospital Medical Center

Cohen Children's Medical Center

CS Mott Children's Hospital/University of Michigan

Dell Children's Medical Center

Hospital for Sick Children, Toronto

Joe DiMaggio Children's Hospital

Nemours, Alfred I. duPont Hospital for Children

Phoenix Children's Hospital

Rady Children's Hospital

Riley Children's Hospital

Seattle Children's Hospital

University of Mississippi

Morgan Stanley Children's Hospital

Medical College of Wisconsin, Children's Hospital of Wisconsin

Ann & Robert Lurie Children's Hospital Chicago

Valley Children's Healthcare and Hospital

UT Southwestern Medical Center, Children's Health Dallas

University of Alabama

University of North Carolina

**Rutgers Robert Wood Johnson Medical School**

Rutgers Robert Wood Johnson Medical School

Baylor College of Medicine,Texas Children's Hospital

Central Michigan University, Children's Hospital of Michigan

Connecticut Children's Medical Center

Johns Hopkins University

The Regents of the University of California San Francisco

American Academy of Family Physicians, AAFP, NRN, DARTNet Institute

American Academy of Pediatrics

Hackensack Meridian Health Hospitals Corporation

MetroHealth System

New York Medical Center, Westchester Medical Center

Saint Barnabas Medical Center, NBI

Children's Mercy Kansas City

Yale School of Medicine

**University of California San Diego, Main Cohort**

University of California San Diego (Tantisira)

**University of California San Diego, ABCD**

University of California San Diego, ABCD

Children's Hospital, Los Angeles

Florida International University

Laureate Institute for Brain Research

Medical University of South Carolina

Oregon Health & Science University

SRI International

University of California, Los Angeles

University of Colorado Boulder

University of Florida

University of Maryland Baltimore (Pediatric)

University of Michigan

University of Minnesota

University of Pittsburgh Medical Center

University of Rochester

University of Utah

University of Vermont

University of Wisconsin, Milwaukee

Virginia Commonwealth University

Washington University St. Louis

Yale University

**Virginia Commonwealth University, Main Cohort**

Virginia Commonwealth University

NYU Langone Health

Rhode Island Hospital

**Columbia University Irving Medical Center**

Columbia University Irving Medical Center

Best Healthcare Inc.

### **Congenitally Exposed Cohort**

**University of California San Francisco**

University of California San Francisco

**University of Utah**

University of Utah

University of Alabama at Birmingham

University of Texas HSC at Houston, Memorial Hermann Texas Medical Center

University of Texas Medical Branch at Galveston

UH MacDonald's Women's Hospital

Yale University

Wakemed Raleigh/Wakemed North

University of Pittsburgh

University of Pennsylvania

University of North Carolina, Chapel Hill

Saint Peter's University Hospital

Ohio State University

Northwestern University

NorthShore University HealthSystem

New York-Presbyterian, Queens

Miami Valley Hospital

Case Western MetroHealth Medical

Medical College of Wisconsin

Good Samaritan

Duke University Medical Center

ChristianaCare Health

University of Texas HSC at Houston - Memorial City

Brown University Women and Infants Hospital

University of Texas HSC at Houston, LBJ Hospital

Columbia University

### **Autopsy Cohort**

**Brigham and Women's Hospital (BWH)**

Brigham and Women's Hospital (BWH)

Massachusetts General Hospital

**CVPath Institute**

Howard University

NYU Tisch and Winthrop Hospitals

**Mount Sinai**

Mount Sinai Hospital

**University of New Mexico Health Sciences Center**

University of New Mexico Health Sciences Center

**Mayo Clinic**

Mayo Clinic

### **Electronic Health Record (EHR) Cohort**

**ADVANCE**

OCHIN, Inc.

**CAPriCORN**

Northwestern University

Greater Plain Collaborative (GPC)

InterMountain Healthcare

Medical College of Wisconsin

University of Iowa Healthcare

University of Missouri

University of Nebraska Medical Center

University of Utah

UT Southwestern Medical Center

**INSIGHT**

Weill Cornell Medical College

Albert Einstein College of Medicine

Icahn School of Medicine at Mount Sinai

Columbia University Irving Medical Center

New York University School of Medicine

OneFlorida Clinical Research Consortium

University of Florida/UF Health

Emory

USF Tampa

Nicklaus Children's Hospital

University of Miami

**PaTH**

University of Pittsburgh/UPM

Ohio State University

Pennsylvania State University

Temple University

University of Michigan

**PEDSnet**

Children's Hospital of Philadelphia (CHOP)

The Nemours Foundation

Nationwide Children's Hospital

Seattle Children's Hospital

Children’s Hospital Colorado

Ann & Robert H. Lurie Children's Hospital of Chicago

Cincinnati Children’s Hospital Medical Center

Children's National Medical Center

Indiana University

Stanford Medicine Children’s Health

**REACHnet**

Ochsner Health System

University of California San Francisco (UCSF)

University Medical Center New Orleans (LSU) - Institute for Public Health Innovation (IPHI)

Star

Vanderbilt University Medical Center (VUMC)

Duke University

Medical University of South Carolina (MUSC)

Wake Forest Baptist Health
